# Matched liver–serum multi-omics and genetic risk reveal hepatic compartmentalization of metabolic disease in severe obesity

**DOI:** 10.64898/2026.04.16.26351046

**Authors:** Shashank Gupta, Oveis Jamialahmadi, Rosellina M. Mancina, Daniel Duberg, Umberto Vespasiani-Gentilucci, Federica Tavaglione, Vincenzo Bruni, Dario Tuccinardi, Tuulia Hyötyläinen, Stefano Romeo, Matej Orešič

## Abstract

We profiled the hepatic transcriptome, lipidome and metabolome and serum metabolome in 211 MAFALDA participants with severe obesity undergoing bariatric surgery with liver biopsy. In 187 participants with paired samples, liver and serum levels of 40 matched metabolites correlated weakly (median ρ 0.02), and steatosis and lobular inflammation were associated with 116 and 216 of 701 hepatic but ≤1 of 653 serum metabolites. Hepatic amino acids were lower with lobular inflammation without serum changes, independently of tissue fat, dilution and glycemia, but largely shared with steatosis. Serum added little to clinical discrimination of steatosis (area under the curve 0.73 to 0.75) and nothing for inflammation, steatohepatitis or fibrosis. A polygenic score for hepatic lipid retention was associated with higher hepatic but lower serum triglycerides; its effects on inflammation and steatohepatitis were largely mediated by liver fat. Circulating metabolites should be validated against histology, not interpreted as direct measures of hepatic metabolism.

## Main

Metabolic dysfunction-associated steatotic liver disease (MASLD) is the most prevalent chronic liver disease worldwide, affecting over one billion people and representing a leading cause of cirrhosis and hepatocellular carcinoma.^1,2^ MASLD is biologically heterogeneous: people with similar clinical phenotypes follow divergent trajectories from steatosis to steatohepatitis and fibrosis, and the molecular determinants of this heterogeneity remain incompletely understood.^3^

In the circulation, molecular lipid signatures predict liver fat content and discriminate people with and without steatosis, particularly in MASLD accompanied by systemic metabolic dysfunction, and have been developed into non-invasive tests.^4,5^ The liver is a master regulator of circulating lipid and glucose homeostasis.^6^ In this systemic form of MASLD, liver steatosis is accompanied by higher plasma triglycerides through increased very-low-density lipoprotein secretion,^7^ and hepatic lipid quantity and remodeling across the MASLD spectrum are partly mirrored in the serum.^8–11^ This coupling is not universal, however: in liver-specific MASLD, triglycerides are retained in the liver and circulating triglyceride concentrations are lower,^12,13^ so hepatic lipid accumulation does not necessarily translate into higher circulating levels. Moreover, MASLD reduces metabolic adaptability^14^ and is associated with type 2 diabetes, in part through hepatic insulin resistance and increased gluconeogenesis.^15^ It is therefore unclear whether the circulating concentrations of other metabolite classes, such as amino acids, bile acids and purine metabolites, mirror their hepatic concentrations, capture components of the MASLD spectrum beyond steatosis or add diagnostic value to the clinical features already associated with MASLD, such as dyslipidemia and diabetes. Whether hepatic and circulating concentrations of the same metabolite are coupled within an individual, and whether serum adds information beyond routine clinical data, have rarely been tested,^16,17^ although blood-based tests are increasingly used to triage and monitor people with MASLD.

Human genetics offers an independent handle on this question. Variants in genes governing hepatic triglyceride handling, such as *TM6SF2*, raise liver fat while lowering circulating lipids, and genetically raised liver fat promotes liver injury.^18–20^ Partitioned polygenic risk scores (pPRS) separate variants that raise liver fat together with circulating lipids (concordant) from those that retain lipid in the liver while lowering circulating lipids (discordant),^12^ providing a genetic instrument for hepatic–systemic compartmentalization.

In this work, we profiled multi-omics in matched liver tissue and serum from 211 participants of the “Molecular Architecture of Fatty Liver Disease in individuals with obesity undergoing bariatric surgery” (MAFALDA) study,^13,21^ all adults with severe obesity and protocol liver biopsy. We tested whether circulating metabolites mirror the liver concentrations, whether the histological features of MASLD (steatosis, lobular inflammation, metabolic dysfunction-associated steatohepatitis (MASH) and fibrosis) are reflected in the hepatic or the circulating metabolome, and whether serum metabolomics adds discrimination beyond clinical variables. We then used pPRS to test whether genetic hepatic lipid retention separates the two compartments and relates to histological features through liver fat, and evaluated key signatures in three external cohorts.^10,22,23^

## Results

### Cohort characteristics

Among 211 participants with severe obesity undergoing bariatric surgery with protocol liver biopsy, men were older than women (median 48 versus 43 years; *P* = 0.009) with similar body mass index (BMI) (**Table 1**). Men exhibited higher levels of HbA1c, aminotransferases, gamma-glutamyl transferase (GGT), and triglycerides, alongside lower levels of HDL-cholesterol (HDL-C) (all *P* < 0.05). Steatosis and inflammation were more prevalent in men. The proportion with any fibrosis (stage ≥1) was similar in men and women (74% versus 65%; *P* = 0.3), but the distribution of fibrosis stages differed between sexes (*P* = 0.001) and stage ≥2 fibrosis was more frequent in men (40% versus 17%; *P* < 0.001) **(Table 1)**.

**Table 1.** Baseline clinical and histological characteristics of the study cohort, stratified by sex. Participants were adults with severe obesity undergoing bariatric surgery with protocol liver biopsy (n = 211). Histological features were graded using the Kleiner scoring system and are shown both as the full grade/stage distribution and in the dichotomized form used in all downstream analyses (grade/stage ≥1 versus 0). Histologically definite MASH was diagnosed according to the Brunt criteria (steatosis, lobular inflammation and hepatocellular ballooning). Of 211 participants, 139 (66%) had hepatic steatosis of grade ≥1 and therefore met the histological criterion for steatotic liver disease. Lobular inflammation was almost entirely grade 1 (112 of 121 inflammation-positive participants), so the dichotomized inflammation variable reflects the presence of any lobular inflammation rather than a severity gradient. Of the 154 women, 119 (77%) were pre-menopausal and 35 (23%) postmenopausal; menopausal status is not applicable to men and is not tabulated. MASH, metabolic dysfunction-associated steatohepatitis; ALT, alanine aminotransferase; AST, aspartate aminotransferase; GGT, gamma-glutamyl transferase; HDL-C, high-density lipoprotein cholesterol; LDL-C, low-density lipoprotein cholesterol; BMI, body mass index. *Median (IQR); n (%). #*P* values were calculated using the Wilcoxon rank-sum test for continuous variables and the χ^2^ test for categorical variables, without adjustment for multiple comparisons. Bold *P* values denote statistical significance at *P* < 0.05.

| Characteristic | All participants<br>N = 211* | Women<br>N = 154* | Men<br>N = 57* | P<br>value# |
| --- | --- | --- | --- | --- |
| Age, years | 44.0 (35.0, 51.0) | 43.0 (35.0, 49.0) | 48.0 (37.0, 54.0) | 0.009 |
| Body mass index, kg/m<sup>2</sup> | 40.9 (38.0, 44.4) | 40.6 (38.0, 44.0) | 41.6 (38.6, 45.1) | 0.3 |
| Fasting glucose, mg/dL | 99.0 (93.0, 107.0) | 98.0 (92.0, 106.0) | 100.0 (95.0, 111.0) | 0.079 |
| HbA1c, % | 5.6 (5.3, 5.9) | 5.5 (5.3, 5.9) | 5.8 (5.4, 6.1) | 0.002 |
| AST, U/L | 26.0 (22.0, 32.0) | 25.0 (20.0, 30.0) | 31.0 (26.0, 37.0) | <0.001 |
| ALT, U/L | 30.0 (21.0, 42.0) | 26.0 (19.0, 39.0) | 38.0 (29.0, 58.0) | <0.001 |
| GGT, U/L | 24.0 (18.0, 33.0) | 22.0 (17.0, 31.0) | 31.0 (25.0, 41.0) | <0.001 |
| Total cholesterol, mg/dL | 181.0 (163.0, 198.0) | 181.0 (164.0, 204.0) | 177.0 (156.0, 191.0) | 0.083 |
| HDL cholesterol, mg/dL | 46.0 (39.0, 52.0) | 47.0 (42.0, 53.0) | 39.0 (35.0, 45.0) | <0.001 |
| LDL cholesterol, mg/dL | 120.0 (103.0, 143.0) | 122.0 (103.0, 145.0) | 118.0 (104.0, 136.0) | 0.3 |
| Triglycerides, mg/dL | 125.0 (95.0, 164.0) | 122.0 (93.0, 158.0) | 141.0 (97.0, 186.0) | 0.023 |
| Hepatic steatosis grade |  |  |  | <0.001 |
| Grade 0 | 72 (34%) | 65 (42%) | 7 (12%) |  |
| Grade 1 | 71 (34%) | 46 (30%) | 25 (44%) |  |
| Grade 2 | 38 (18%) | 24 (16%) | 14 (25%) |  |
| Grade 3 | 30 (14%) | 19 (12%) | 11 (19%) |  |
| Lobular inflammation grade |  |  |  | 0.002 |
| Grade 0 | 90 (43%) | 77 (50%) | 13 (23%) |  |
| Grade 1 | 112 (53%) | 71 (46%) | 41 (72%) |  |
| Grade 2 | 9 (4.3%) | 6 (3.9%) | 3 (5.3%) |  |
| Fibrosis stage |  |  |  | 0.001 |
| Stage 0 | 69 (33%) | 54 (35%) | 15 (26%) |  |
| Stage 1 | 93 (44%) | 74 (48%) | 19 (33%) |  |
| Stage 2 | 44 (21%) | 25 (16%) | 19 (33%) |  |
| Stage 3 | 5 (2.4%) | 1 (0.6%) | 4 (7.0%) |  |
| Histologically definite MASH | 87 (41%) | 52 (34%) | 35 (61%) | <0.001 |
| Steatosis ≥ grade 1 | 139 (66%) | 89 (58%) | 50 (88%) | <0.001 |
| Lobular inflammation ≥ grade 1 | 121 (57%) | 77 (50%) | 44 (77%) | <0.001 |
| Fibrosis ≥ stage 1 | 142 (67%) | 100 (65%) | 42 (74%) | 0.3 |

### Transcriptomic shifts reflect stage-specific metabolic and inflammatory programs

As expected, sex had a strong effect on global liver gene expression (**Fig. S1**), and all models were adjusted for sex. Among 208 participants with liver RNA-seq, 1,532 genes were differentially expressed with steatosis (987 up, 545 down), 1,852 with lobular inflammation (1,094 up, 758 down) and 93 with fibrosis (65 up, 28 down) (false discovery rate [FDR] ≤ 0.05; **Fig. 1a, Table S1**). Fibrosis was of low to moderate degree in the MAFALDA cohort, which probably explains the smaller fibrosis signature. Steatosis and inflammation shared most differentially expressed genes (DEGs) (727 up, 457 down), whereas few genes were specific to fibrosis (12 up, 10 down; **Fig. 1b, Table S2a**). Downregulated genes in steatosis and inflammation were enriched for complement and coagulation cascades and amino acid metabolism, and upregulated genes for ECM–receptor interaction and focal adhesion (**Fig. 1c, Table S2b**). Interestingly, the fibrosis signature comprised interferon-stimulated genes (*OAS2, MX1, ISG15, IFI27*), as well as cell-cycle and matrix genes (*CDKN1A, CDK6, PDGFRB, COL1A1*),^22,24^ with lower expression of tryptophan metabolism genes.

**Fig. 1.**
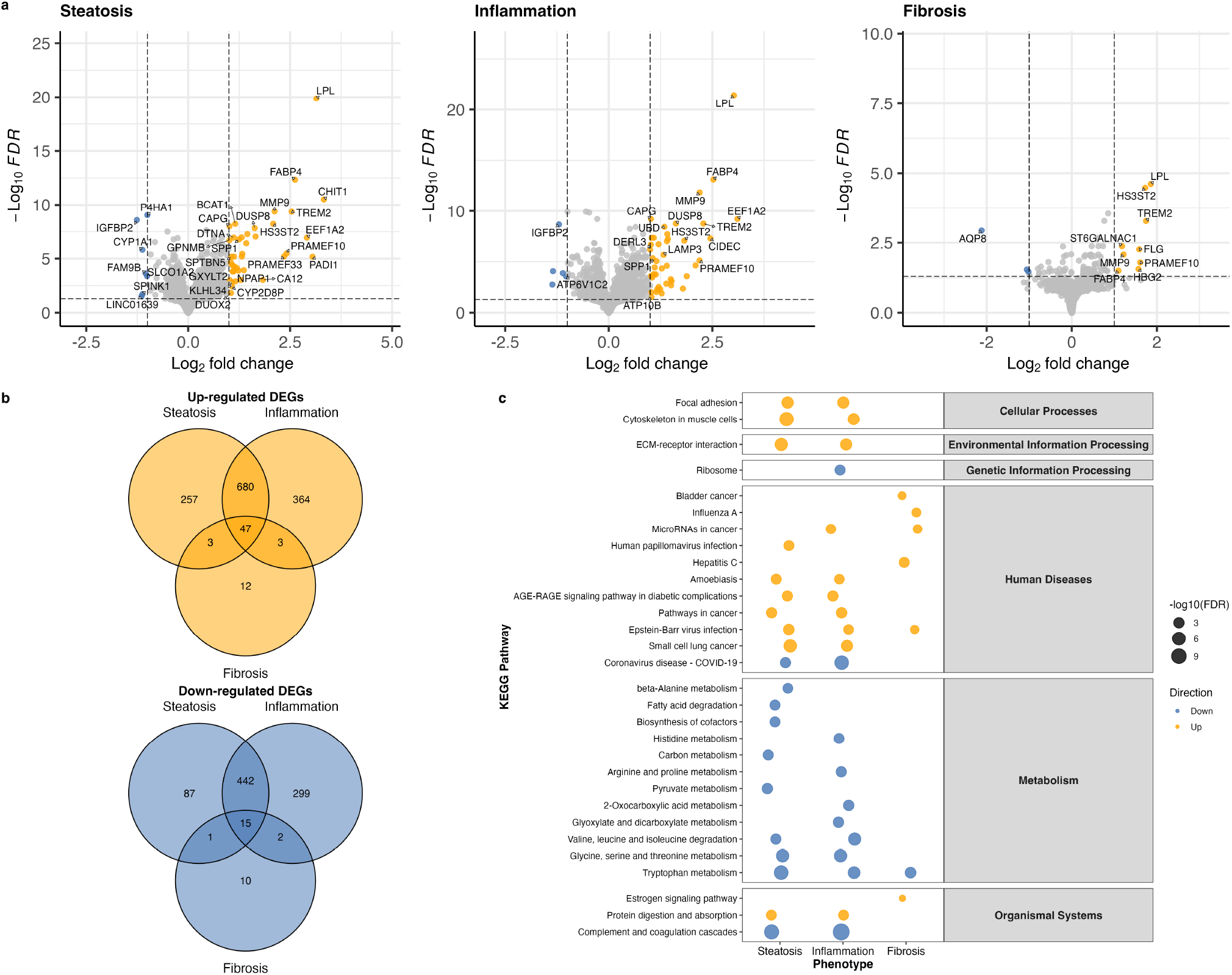
Differentially expressed genes (DEGs) across MASLD histological features. Liver RNA-seq in 208 participants; DESeq2 models adjusted for age, BMI and sex; DEGs at FDR ≤ 0.05. **(a)** Volcano plots of genes associated with steatosis (1,532 DEGs), lobular inflammation (1,852) and fibrosis (93); orange and blue, up- and downregulated DEGs with |log2 fold change| > 1; gray, other genes; dashed lines, FDR = 0.05 and |log2 fold change| = 1; selected top-ranked genes are labeled. **(b)** Overlap of upregulated (top) and downregulated (bottom) DEGs across the three features. (c) KEGG pathways enriched among up- and downregulated DEGs (FDR ≤ 0.10, ≥ 3 genes; background, all tested genes).

### Hepatic lipidomic and metabolomic signatures

Hepatic lipidomics and polar metabolomics showed extensive differences (**Fig. 2, Tables S3–S4**). Lobular inflammation was associated with lower hepatic amino acids and higher bile acids, and both steatosis and inflammation with lower polyunsaturated fatty acids (**Table S4**), accumulation of triglycerides and depletion of phosphatidylcholines, sphingomyelins and ceramides (**Table S3**). Joint pathway analysis of hepatic genes and metabolites highlighted differences in glycolysis, linoleic acid and tryptophan metabolism (**Fig. S2**).

**Fig. 2.**
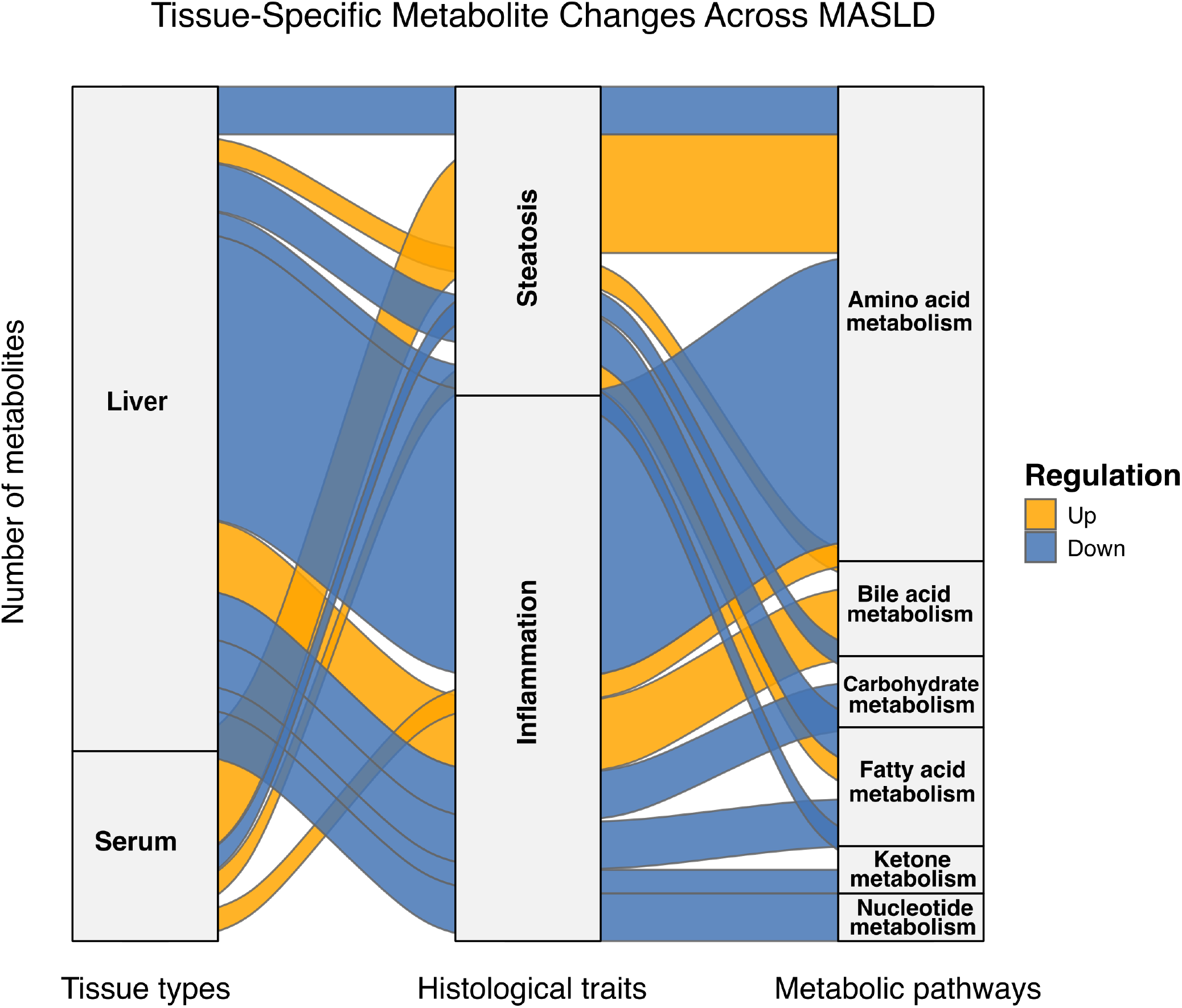
Differential metabolites in liver and serum. Sankey diagram of metabolites associated with steatosis and lobular inflammation (FDR ≤ 0.10; limma adjusted for age, sex and BMI) in liver (n = 189) and serum (n = 209), grouped by pathway class; stream width, number of unique metabolites; orange, higher; blue, lower. Unclassified and putative metabolites are omitted. Hepatic amino acids were mostly lower and serum amino acids mostly higher with steatosis and lobular inflammation (grade ≥1 versus 0), a divergence tested directly in the matched subset (Fig. 4).

### External validation

To test whether the MAFALDA results also apply to people without severe obesity, we compared our signatures with two independent liver transcriptomic cohorts spanning a wider range of BMI, GSE135251 and GSE162694.^22,23^ Of the 1,532 steatosis DEGs, 683 were also significant in GSE162694 and 853 in GSE135251; of the 1,852 inflammation DEGs, 761 and 969; and of the 93 fibrosis DEGs, 46 and 49, respectively (**Fig. 3a–b, Table S5**). Of these shared genes, 97.8–99.4% changed in the same direction (log2 fold-change ρ 0.67–0.74 for fibrosis and 0.87–0.89 for steatosis and inflammation; **Fig. 3c**), *KRT10* being the only discordant fibrosis gene. Replication rates of discovery DEGs were 48–52% for fibrosis, 44–55% for steatosis and 41–52% for inflammation, and results were similar after adjustment for inferred sex in GSE135251 (**Tables S5–S6**). Six of seven discovery fibrosis Kyoto Encyclopedia of Genes and Genomes (KEGG) pathways were enriched in at least one external cohort (**Fig. 3d, Table S7**). In the ST000915 liver lipidomic cohort (29 normal, 17 steatosis), 48 of 50 triglyceride measurements matched to discovery steatosis features were directionally concordant, 47 of them significantly, whereas 13 of 14 phosphatidylcholine measurements went in the opposite direction (**Tables S8–S9**). Taken together, the transcriptomic signatures and the hepatic triglyceride signature were partly reproduced in cohorts spanning a wider BMI range, whereas the phosphatidylcholine signature was not.

**Fig. 3.**
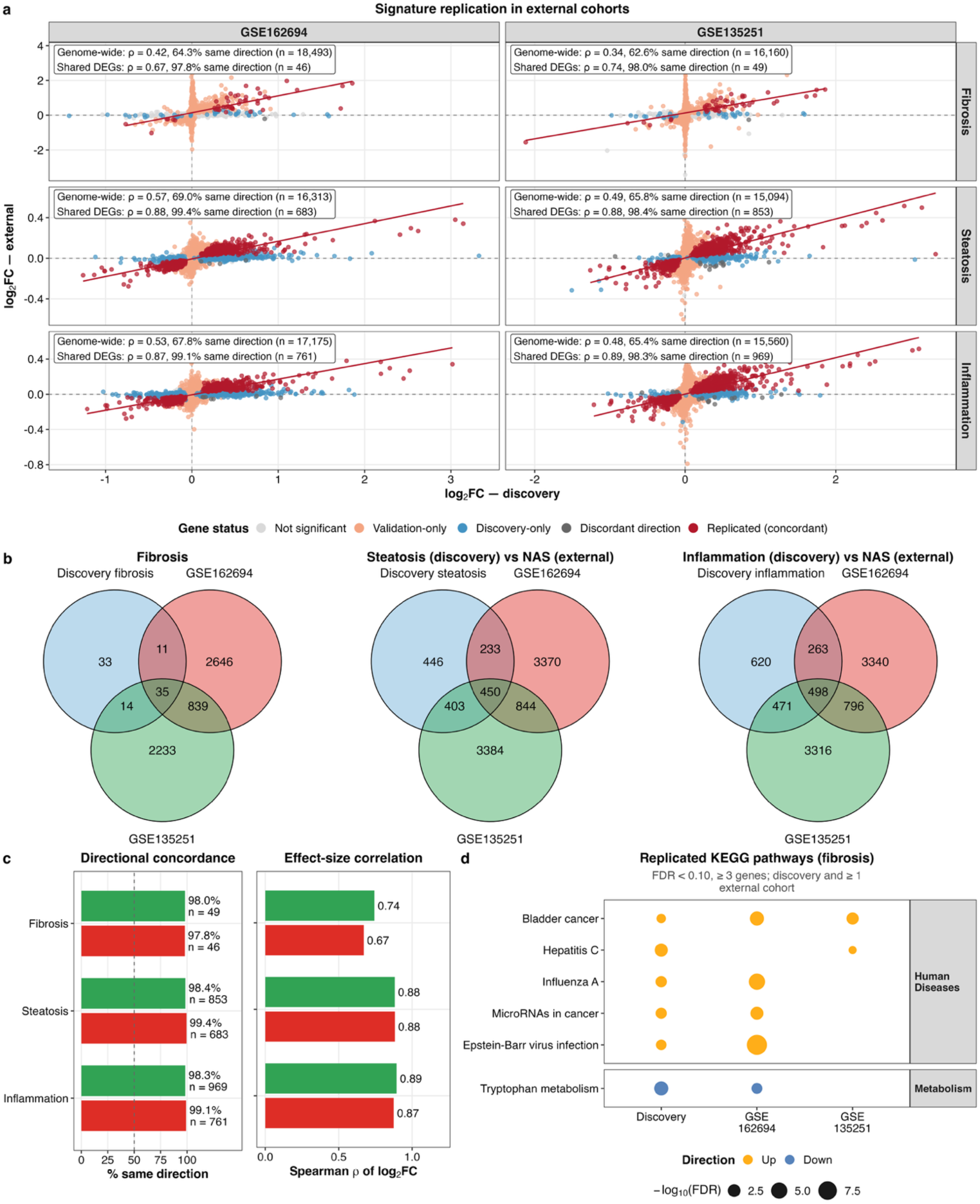
External validation of hepatic transcriptomic signatures. Discovery: 208 participants (93 DEGs associated with fibrosis, 1,532 with steatosis and 1,852 with inflammation; FDR ≤ 0.05). External cohorts: GSE162694 and GSE135251; fibrosis tested as F0 versus F1–F4, steatosis and inflammation against the NAS. (a) Log2 fold changes in the discovery versus each external cohort for all genes measured in both; red, significant in both in the same direction; dark gray, significant in both in opposite directions; blue, discovery only; salmon, external only; light gray, neither; the line is fitted to shared DEGs. (b) Overlap of DEGs between cohorts. (c) Percentage of shared DEGs changing in the same direction (dashed line, 50%) and Spearman correlation of their log2 fold changes. (d) KEGG pathways enriched among fibrosis DEGs (FDR ≤ 0.10, ≥3 genes) in the discovery cohort and at least one external cohort; orange, upregulated; blue, downregulated; point size, −log10 FDR.

### Circulating metabolome does not mirror the liver

In serum, 12 metabolites were associated with steatosis and four with lobular inflammation (FDR ≤ 0.10; 13 unique features, five unannotated; **Fig. 2, Table S10a**); leucine/isoleucine was the strongest annotated association with inflammation and glutamic acid with steatosis. In the 187 participants with matched samples, hepatic and serum concentrations of the 40 metabolites measured in both compartments were weakly correlated (median Spearman ρ 0.02, interquartile range −0.06 to 0.14; 9 of 40 at FDR ≤ 0.10; **Fig. 4a, Table S10b**); as expected for positive controls, the strongest liver–serum correlations were for two metabolites derived from exogenous precursors, the gut microbial metabolite p-cresol sulfate (ρ = 0.51) and the paracetamol metabolite 4-acetamidophenol sulfate (ρ = 0.29).

**Fig. 4.**
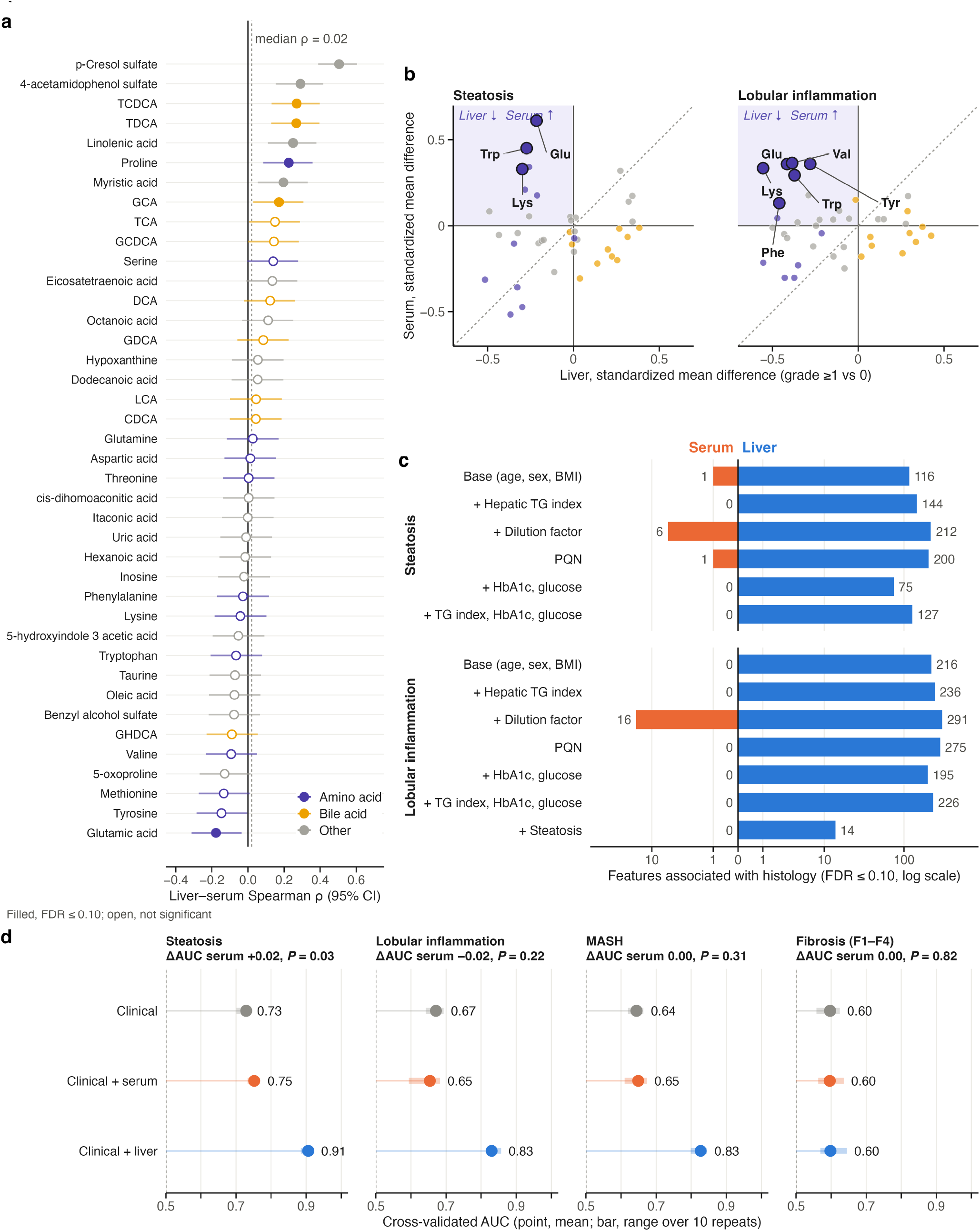
Hepatic versus circulating metabolome in matched samples (n = 187). (a) Liver–serum Spearman correlations for all 40 matched metabolites, ranked, with Fisher 95% confidence intervals; filled points, FDR ≤ 0.10; dashed line, median ρ; colors indicate metabolite class. (b) Association with the presence of steatosis or lobular inflammation (grade ≥1 versus 0) in liver (x) and serum (y) (standardized mean difference; limma adjusted for age, sex and BMI; Table S11b). Outlined points, labeled with three-letter amino-acid codes, differ significantly between compartments (FDR ≤ 0.10); the shaded quadrant marks metabolites lower in liver but higher in serum; the dashed line is the identity. (c) Numbers of histology-associated features (FDR ≤ 0.10) in liver (right) and serum (left) under the base model and after adjustment for hepatic triglyceride content, a sample dilution factor, probabilistic quotient normalization, HbA1c and glucose, the triglyceride index with HbA1c and glucose, or steatosis (log scale). (d) Cross-validated discrimination (elastic net, 10 × 5-fold) by clinical variables (age, sex, BMI, HbA1c, glucose) alone or with serum or hepatic metabolites; points, mean area under the receiver operating characteristic curve (AUC); bars, range over repeats; the AUC gain from serum and its DeLong *P* value are given above each outcome.

### Hepatic but not serum metabolome predicts liver histological features

In the 187 participants with matched samples, 116 of 701 hepatic metabolites were associated with steatosis and 216 with lobular inflammation, compared with 1 and 0 of 653 serum metabolites (**Table S11a**) and 12 and 4 in all 209 participants with serum data (**Table S10a**). On the matched panel of 40 metabolites, effect sizes were larger in liver than in serum for inflammation (median absolute standardized difference 0.29 versus 0.11; *P* = 5 × 10^−6^) but not significantly different for steatosis (0.26 versus 0.12; *P* = 0.19) (**Table S11b**).

In the paired liver–serum contrasts (limma on the within-subject difference, FDR ≤ 0.10), three metabolites differed significantly between compartments for steatosis (glutamic acid, tryptophan and lysine) and six for lobular inflammation (lysine, glutamic acid, valine, tryptophan, tyrosine and phenylalanine); all were amino acids (six distinct), and all were lower in liver, with unchanged or higher serum levels (**Fig. 4b, Tables S10b** and **S11c**). The discordance arose in the liver. For lobular inflammation, all twelve matched amino acids had lower hepatic levels, significantly so for ten, whereas none changed significantly in serum; five of the six discordant amino acids were themselves significant in liver, tyrosine being the exception (**Table S11c**). The same direction was present for steatosis — eleven of twelve amino acids lower in liver — but only aspartic acid reached significance, consistent with the smaller compartment difference for that phenotype.

Bile acids showed the opposite pattern, being higher in liver and lower in serum, but only as a directional trend: for lobular inflammation nine of ten matched bile acids had opposite liver–serum directions (two-sided sign test *P* = 0.021; **Table S10b**, which also gives the tests for amino acids and for steatosis) and eight of ten were higher in liver and lower in serum, yet just three (glycocholic, taurocholic and taurodeoxycholic acid) were significantly raised in liver and none reached FDR ≤ 0.10 in the paired contrast. Bile acids were also among the few metabolites whose hepatic and serum levels tracked one another within an individual (taurochenodeoxycholic acid ρ = 0.27, taurodeoxycholic acid ρ = 0.27; both FDR ≤ 0.01), indicating that the liver–serum uncoupling seen for amino acids is not uniform across metabolite classes.

Adjustment for hepatic triglyceride content or a sample dilution factor, or probabilistic quotient normalization, increased rather than reduced the number of hepatic associations (144–291), and all six amino acids remained discordant for inflammation, indicating that hepatic amino acid depletion is not a dilution effect of steatosis (**Fig. 4c, Table S11a**). Additional adjustment for HbA1c and glucose left the amino acid discordance for inflammation unchanged, although hepatic associations with steatosis fell from 116 to 75, and serum associations remained few (0–16) across these models. After adjustment for steatosis, however, hepatic associations with inflammation fell from 216 to 14 and only lysine and valine remained discordant, suggesting that most inflammation associations were driven by steatosis in the base model, although, when hepatic triglyceride content was also modeled as a continuous index, about half of the inflammation associations persisted after steatosis adjustment (113 of 236). Excluding hepatic features correlated with the triglyceride index (|ρ| > 0.5) reduced these counts by 1–13% in models not adjusted for steatosis (65–288 features; **Table S11a**), indicating that the hepatic signal is not an artifact of tissue lipid content. Adding serum metabolites to clinical variables produced a modest and not clinically meaningful improvement in the discrimination of steatosis (area under the receiver operating characteristic curve [AUC] 0.73 to 0.75; *P* = 0.03) and none for inflammation, MASH or fibrosis; serum metabolites alone discriminated steatosis at least as well as clinical variables (AUC 0.78 versus 0.73), although combining the two did not improve on serum alone (0.75; **Table S11d**). Adding hepatic metabolites instead improved discrimination substantially, from 0.73 to 0.91 for steatosis, from 0.67 to 0.83 for lobular inflammation and from 0.64 to 0.83 for MASH (all *P* < 10^−5^), but not for fibrosis (0.60 to 0.60; *P* = 0.58) (**Fig. 4d, Table S11d–e**).

### Genetic hepatic lipid retention separates hepatic and circulating triglycerides

In all 211 participants, the discordant and concordant pPRS were not correlated (r = −0.04) and independent of age, sex or BMI (all *P* > 0.39). The discordant pPRS was robustly associated with higher hepatic triglyceride content (β = 0.33 SD per SD, *P* = 2.6 × 10^−7^) but lower serum triglycerides (β = −0.22, *P* = 8.2 × 10^−4^), and with steatosis grade (β = 0.28, *P* = 5.0 × 10^−5^) and NAFLD activity score (NAS; β = 0.40, *P* = 7.7 × 10^−4^); the concordant pPRS was associated with neither hepatic nor serum triglycerides (*P* ≥ 0.40) and only weakly with steatosis (β = 0.14, *P* = 0.049) (**Fig. 5a–b, Table S12a**).

**Fig. 5.**
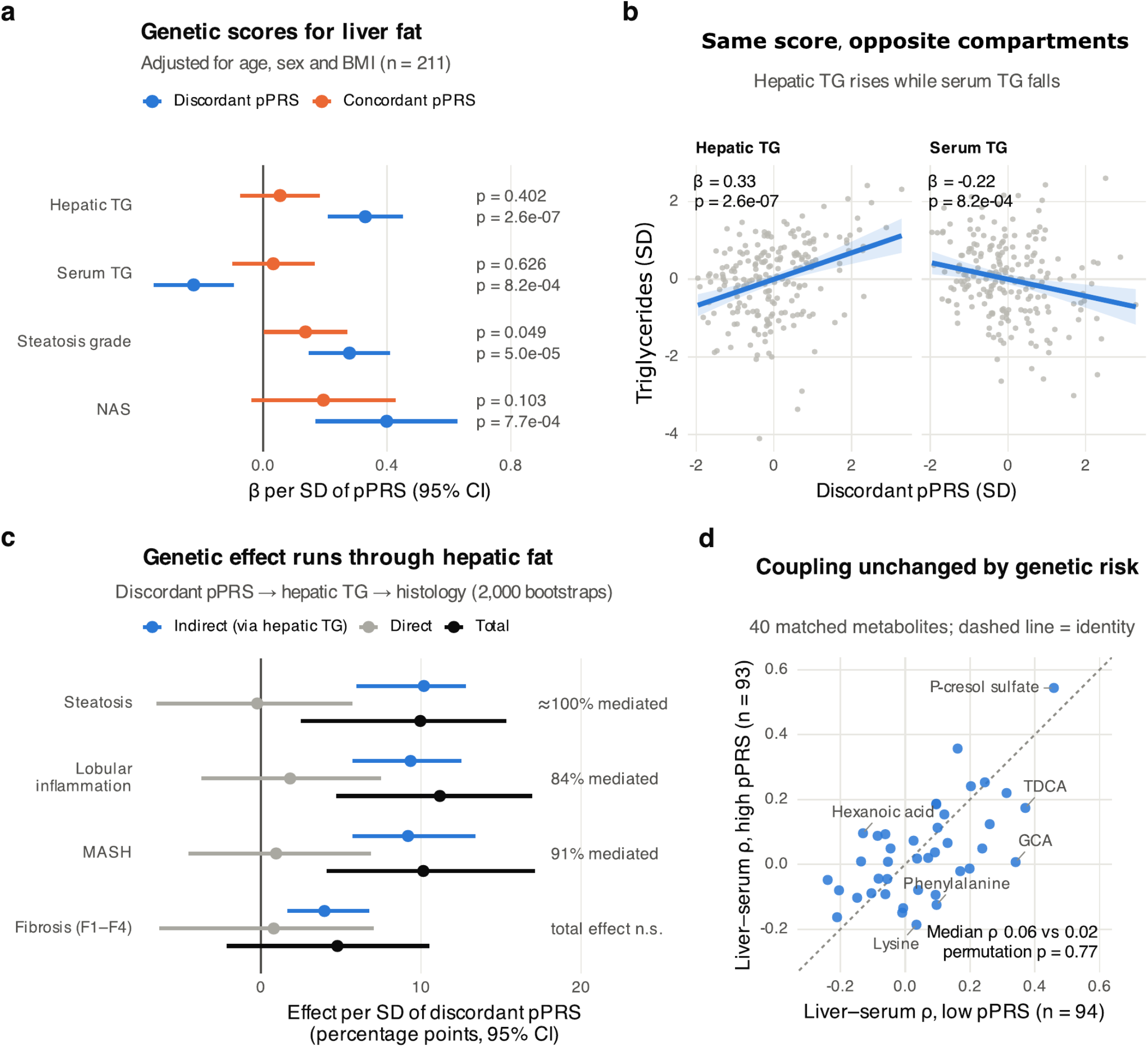
Genetic predisposition to hepatic lipid retention. **(a)** Associations of the discordant or the concordant partitioned polygenic risk scores (pPRS) with the hepatic triglyceride (TG) index, log-transformed serum TG (both standardized), steatosis grade and NAS (β per SD of pPRS with 95% CI; linear regression adjusted for age, sex and BMI; n = 211; *P* values shown on the right; Table S12a). **(b)** Discordant pPRS versus standardized hepatic and serum TG; lines, linear fits with 95% confidence bands; β and *P* from the models in **a. (c)** Mediation of discordant pPRS effects on steatosis, lobular inflammation, MASH and any fibrosis (F1–F4 versus F0) through hepatic TG content (linear mediator and logistic outcome models adjusted for age, sex and BMI; n = 211): indirect (*via* hepatic TG), direct and total effects in percentage points per SD (95% bootstrap CI, 2,000 resamples; Table S12b). Labels give the proportion mediated; for steatosis the estimate was 102% (direct effect ≈ 0), shown as ≈100% (complete mediation); n.s., total effect not significant. **(d) Liver–serum Spearman** correlations of the 40 matched metabolites in participants below (n = 94) and above (n = 93) the median discordant pPRS; dashed line, identity; selected metabolites are labeled; median ρ and permutation *P* (2,000 permutations) are shown (Table S12d).

The associations of the discordant pPRS with steatosis, lobular inflammation and MASH were largely mediated by hepatic triglyceride content (indirect effects 10.2 [95% CI 6.0–12.8], 9.4 [5.7–12.5] and 9.2 [5.7–13.4] percentage points per SD; proportion mediated 102% (effectively complete), 84% and 91%, respectively; direct effects *P* ≥ 0.53), whereas no mediation was observed for the concordant pPRS (all FDR = 0.45; **Fig. 5c, Table S12b**). For any fibrosis, the indirect effect was 4.0 percentage points per SD (95% CI 1.6–6.8), but the total effect was not significant (4.8, *P* = 0.15). Adjustment for both scores left the amino acid discordance unchanged (six of six for inflammation), although serum associations rose from 1 to 30 of 653 for steatosis and from none to four for inflammation (**Table S12c**). The discordant pPRS did not modify liver–serum correlations of the 40 matched metabolites (median ρ 0.06 versus 0.02 below and above the median; permutation *P* = 0.77; **Fig. 5d, Table S12d**), the coupling of hepatic with serum triglycerides (interaction *P* = 0.54; **Table S12e**), the within-participant liver–serum difference of any matched metabolite (all FDR ≥ 0.95; **Table S12f**) or serum discrimination of histology (**Table S12g**).

### Selected hepatic amino-acid transport and catabolic genes are transcriptionally reduced

To place the compartment differences in a pathway context, we mapped hepatic transcripts, lipids and metabolites and their serum counterparts onto the main liver–blood exchange pathways (**Fig. 6**). Several plasma-membrane amino acid transporter genes were downregulated in grade ≥1 versus grade 0: *SLC3A2* (log2 fold change −0.17 for steatosis and −0.19 for lobular inflammation), which encodes the heavy chain of heterodimeric amino acid transporters including LAT1, the cationic amino acid transporter *SLC7A2* (CAT2; −0.21 for both) and, for steatosis, the glutamate transporter *SLC1A2* (EAAT2; −0.39) (**Table S1**). Others were higher, notably *SLC1A3* (EAAT1; 0.50 and 0.49), *SLC7A1* (CAT1; 0.35 for inflammation) and *SLC7A6* (0.25 and 0.26), which may partly reflect non-parenchymal-cell expression. Several amino acid catabolic genes were downregulated, including branched-chain (*IVD, ACADSB, ALDH6A1, MMUT*; −0.14 to −0.36), tryptophan–kynurenine (*KYNU, HAAO, ACMSD, AADAT*; −0.15 to −0.28), methionine-cycle (*MAT1A, GNMT, AHCY*; −0.22 to −0.52) and transamination genes (*GOT1, GPT2*; −0.21 to −0.32). Each of the six amino acids that differed between liver and serum is a substrate of at least one of these transporters or pathways. *BCAT1* was higher (1.15 and 1.04), together with markers of lipid-associated macrophages (*TREM2, GPNMB, SPP1*), consistent with a contribution of non-parenchymal cells to the bulk transcriptome. For bile acids, whose hepatic associations were mostly positive and serum associations near zero or negative, expression of the canalicular export pump *ABCB11* (BSEP) was lower (−0.22 and −0.18), whereas the basolateral exporter *ABCC4* (MRP4) and *CYP7A1* were higher with inflammation (0.33 and 0.42). Lower hepatic expression of amino acid catabolic genes, together with lower expression of these transporters, is consistent with altered hepatic amino acid handling, although transcript levels do not measure flux.

**Fig. 6.**
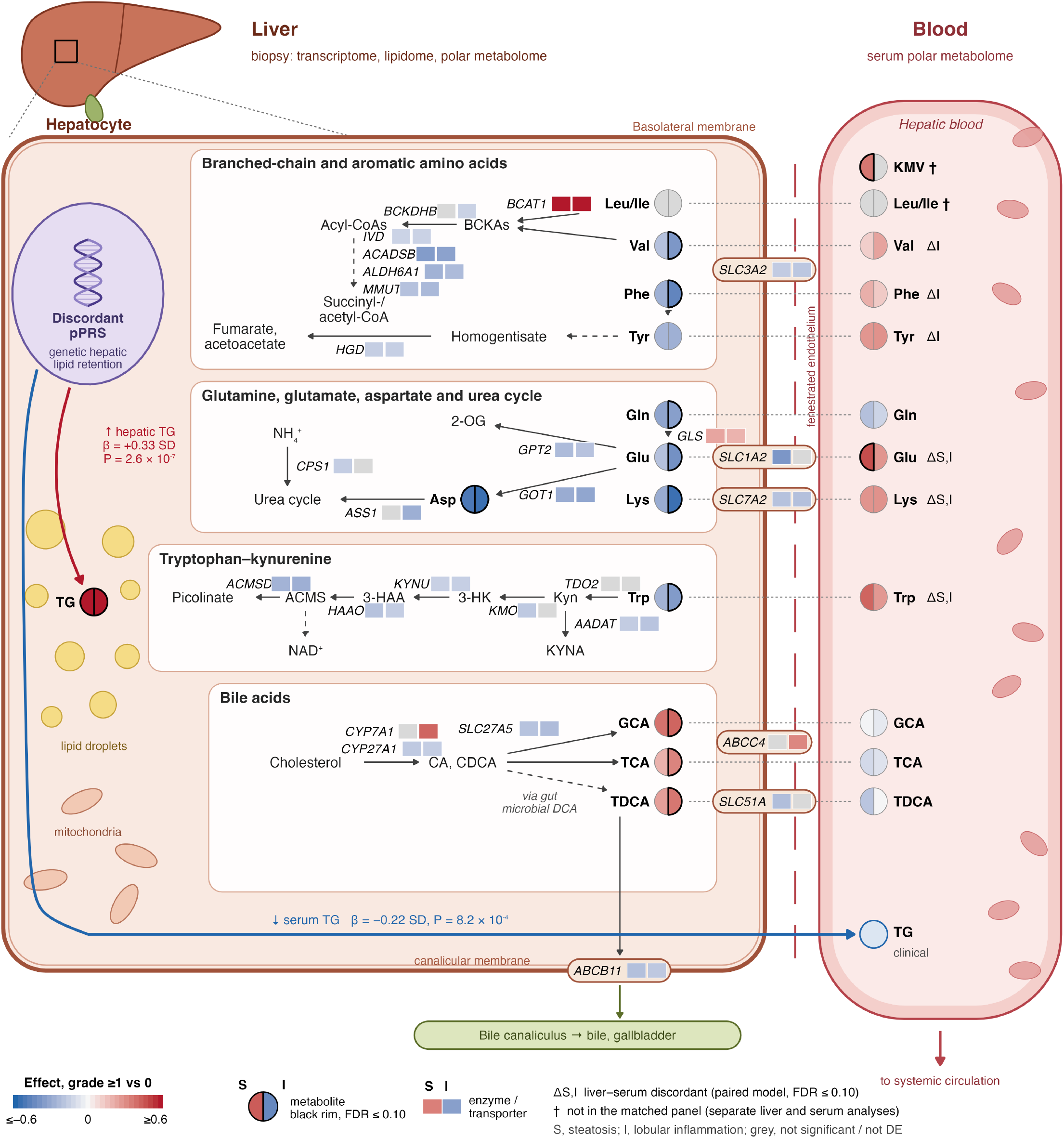
Liver–serum view of hepatic amino acid, tryptophan and bile acid pathways in severe obesity. Schematic of a hepatocyte (zoomed from the liver) facing the hepatic blood. Pathway genes (italic) carry two tiles, steatosis (left) and lobular inflammation (right): hepatic RNA-seq (n = 208), DESeq2 apeglm-shrunk log2 fold change, grade ≥1 versus 0; gray, not differentially expressed (FDR > 0.05; Table S1). Transporters are drawn on the basolateral or canalicular membrane; they were selected from the differentially expressed genes and are not an exhaustive map of hepatic transport. SLC3A2 is the heavy chain of heterodimeric amino-acid transporters; its neutral-amino-acid light-chain partners SLC7A5 (LAT1) and SLC7A8 (LAT2) were not differentially expressed. SLC51A (OSTα) functions with SLC51B (OSTβ), which was not differentially expressed. Other amino-acid transporters were higher: SLC1A3 (steatosis 0.50, inflammation 0.49), SLC7A1 (inflammation 0.35) and SLC7A6 (0.25, 0.26). Circles show metabolites in liver (left of the membrane) and serum (blood): left half, steatosis; right half, lobular inflammation; fill, standardized effect (SD units; limma adjusted for age, sex and BMI); black rim, FDR ≤ 0.10. Liver and serum values are from the matched panel (n = 187; Table S10b); Δ, liver–serum discordant (phenotype effect on the within-participant liver-minus-serum difference, FDR ≤ 0.10). †, not in the matched panel: leucine/isoleucine (one unresolved feature, m/z 130.087) and 3-methyl-2-oxovalerate (KMV), from the separate hepatic (n = 189; Table S4) and serum (n = 209; Table S10a) analyses, significant features only. Hepatic TG, median effect of significant triglyceride species (Table S3). The discordant partitioned polygenic risk score (pPRS) was associated with higher hepatic and lower clinical serum triglycerides (β per SD; n = 211; Table S12a); the serum TG circle shows this genetic association, not a histology effect. Arrows show pathway topology, not measured flux; dashed arrows, several steps. TDCA is formed from deoxycholic acid produced by gut bacteria and conjugated in the liver. Gene expression is from bulk tissue and includes non-parenchymal cells; BCAT1 and GLS (rather than the hepatocyte isoform GLS2) may partly reflect lipid-associated macrophages and other non-parenchymal cells. 2-OG, 2-oxoglutarate; 3-HAA, 3-hydroxyanthranilate; 3-HK, 3-hydroxykynurenine; ACMS, 2-amino-3-carboxymuconate semialdehyde; BCKA, branched-chain 2-oxo acid; CA, cholic acid; CDCA, chenodeoxycholic acid; DCA, deoxycholic acid; GCA, glycocholic acid; KMV, 3-methyl-2-oxovalerate; KYNA, kynurenic acid; Kyn, kynurenine; NAD^+^, nicotinamide adenine dinucleotide; TCA, taurocholic acid; TDCA, taurodeoxycholic acid; TG, triglycerides.

### Exploratory sex-stratified networks and mediation

Hepatic lipid modules showed the strongest associations with histology in both women and men (|ρ| up to 0.77). No module–histology association differed significantly between sexes (0 of 231 at FDR ≤ 0.10), and the larger number of associations in women (51 versus 21) was consistent with the difference in sample size (size-matched two-sided *P* = 0.82). Statistical mediation linked transcriptomic modules to steatosis and inflammation through hepatic lipid modules in women whereas no pathway reached significance in men, in whom 20 of 42 candidate models could not be estimated. These hypothesis-generating results are given in **Figs. S3–S4** and **Tables S13–S16**.

## Discussion

The main findings of this study are that a) serum does not mirror the liver metabolome in people living with severe obesity; b) liver, but not serum, metabolome meaningfully increases the accuracy of clinical variables for diagnosing steatosis, lobular inflammation and MASH, but not fibrosis; and c) genetic predisposition to hepatic lipid retention was associated with higher hepatic but lower circulating triglycerides, and its association with inflammation and MASH was largely mediated by hepatic triglyceride content (summarized in **Fig. S5**).

Amino acids were compartmentalized in liver with inflammation without a corresponding serum change, largely reflecting their shared association with steatosis, whereas bile acids tended to increase in liver without a serum increase. This agrees with reports that serum polar metabolites track histology poorly,^25^ contrasts with the closer liver–serum agreement reported for lipids,^10,11^ and supports compartment-specific metabolic information.^5^ Lower hepatic expression of some amino acid transporters (*SLC3A2, SLC7A2, SLC1A2*) and of amino acid catabolic enzymes may contribute to the observed compartmentalization: reduced hepatic uptake would lower hepatic amino acid concentrations while leaving circulating concentrations unchanged or higher, and lower expression of catabolic enzymes may reflect a reduced amino acid supply to the liver. Similarly, lower expression of the canalicular bile salt export pump (*ABCB11*) is consistent with hepatic bile acid retention, whereas higher basolateral *ABCC4* may represent an adaptive efflux route. These transcript-level associations require confirmation by flux or tracer studies. The amino acid compartmentalization was maintained after adjustment for tissue fat content, sample dilution, glycemia and genetic risk, and liver–serum differences were metabolite-specific rather than global. These findings do not preclude the clinical utility of circulating biomarkers, as serum metabolite concentrations reflect hepatic production and uptake, interorgan exchange, renal clearance, and dietary intake. Moreover, our analysis assessed static concentrations of metabolites measurable in both serum and liver, rather than metabolic flux. The conclusion supported by these data is therefore that the serum concentration of an individual metabolite cannot reliably predict its hepatic concentration.

The genetic analysis provides an independent line of evidence for compartmentalization, in this case of liver triglyceride content. A polygenic score for variants that raise liver fat while lowering circulating lipids, which include *PNPLA3* and *TM6SF2* variants,^12^ was associated with higher hepatic but lower serum triglycerides, and its association with inflammation and MASH was largely accounted for by hepatic triglyceride content. This supports Mendelian randomization evidence that genetically raised liver fat promotes liver injury^19^ and extends the partitioned polygenic risk score approach, which separates variants that raise liver fat together with circulating lipids from those that retain lipid in the liver while lowering circulating lipids,^12^ to direct measurements in liver tissue; because genetic variants are allocated at conception and precede disease, these results are less likely to reflect reverse causation than mediation among simultaneously measured omics layers. By contrast, genetic risk did not modify the weak correlation between hepatic and circulating polar metabolite concentrations, supporting the interpretation that physiological regulation, rather than genetically determined hepatic retention, governs their circulating levels. These data have at least two implications. First, blood metabolite panels should be developed and validated as empirical classifiers of histology rather than as read-outs of hepatic metabolism; in our data, serum did not add clinically meaningful diagnostic accuracy beyond clinical variables. Second, because discordant variants raise liver fat while lowering circulating triglycerides, serum lipid-based markers may underestimate hepatic fat in carriers.

Strengths of the study include the presence of liver histology in every participant, matched liver and serum sampling, four omics layers and genetic risk scores in one cohort, and external validation of transcriptomic and lipidomic signatures.

This study has limitations. First, the cross-sectional design means that mediation estimates rely on the assumed sequence of events and on there being no unmeasured factors affecting both the mediator and the outcome. Second, external validation varied by layer: fibrosis was compared directly, but steatosis and inflammation were compared with the NAS, and the ST000915 comparison differed from the discovery cohort in phenotype definition and feature coverage. Third, only 40 metabolites could be matched between compartments and leucine and isoleucine could not be separated. Fourth, gene expression was measured in whole liver tissue, so some signals, including *BCAT1* and the macrophage markers *TREM2, GPNMB* and *SPP1*, may reflect lipid-associated macrophages rather than hepatocyte metabolism.

In conclusion, in adults with severe obesity, the serum metabolome did not reflect the hepatic metabolome. Histological features were more strongly associated with hepatic than circulating metabolite concentrations, and serum metabolites provided limited diagnostic information beyond routine clinical variables. Genetically determined hepatic lipid retention was associated with higher hepatic and lower circulating triglyceride concentrations, and mediation analyses supported a role for hepatic triglycerides in its association with inflammation and MASH. These findings indicate that circulating metabolite concentrations should be interpreted cautiously as markers of hepatic metabolism.

## Methods

### Study design and participants

We studied 211 adults with severe obesity (154 women, 57 men) from the MAFALDA study,^13,21^ recruited consecutively at bariatric surgery between May 2020 and June 2021 at Fondazione Policlinico Universitario Campus Bio-Medico, Rome, Italy. Liver biopsy and serum were collected in the same operative session. Laparoscopic-guided percutaneous core liver biopsies were scored by pathologists blinded to clinical data for steatosis (0–3), lobular inflammation (0–3), hepatocellular ballooning (0–2) and fibrosis (0–4) according to the Kleiner system, and the NAFLD activity score (NAS) was calculated; histologically definite MASH was diagnosed according to the Brunt criteria (steatosis, lobular inflammation and hepatocellular ballooning). Steatosis, lobular inflammation and fibrosis were dichotomized as grade or stage 0 versus ≥1. Sex was taken from clinical records and included as a covariate in all pooled analyses. The study was approved by the Local Research Ethics Committee at Campus Bio-Medico University Hospital (no. 16/20) and the Swedish Ethics Review Authority (Dnr 2025-08073-01) and followed the Declaration of Helsinki; all participants gave written informed consent.

### Omics profiling

Hepatic lipids and hepatic and serum polar/semipolar metabolites were extracted separately from randomized samples and analyzed by ultra-high-performance liquid chromatography–quadrupole time-of-flight mass spectrometry (UHPLC-QTOFMS) with internal standards and pooled quality-control samples,^9,26^ and raw data were converted to mzML with ProteoWizard^27^ and processed in MZmine 4.3.^28^ Hepatic lipidomics, hepatic polar metabolomics and serum polar metabolomics were available for 211, 189 and 209 participants, respectively. Liver RNA-seq data were available for 208 participants (153 women, 55 men); reads were aligned with STAR and quantified with RSEM;^29,30^ genes with more than 10 reads in total were retained for variance-stabilized analyses (34,852 genes), and genes with ≥10 reads in ≥10 samples for differential expression (21,114 genes). Protocols, parameters and quality control are given in the Supplementary methods and **Table S17**.

### Differential analyses

Analyses were performed in R 4.4.1. Lipid and metabolite features (half-minimum imputation, log2 transformation and autoscaling^31^) were tested with limma^32^ and gene expression with DESeq2^33^ (Wald test with independent filtering at α = 0.05, apeglm shrinkage^34^), adjusting for age, sex and BMI. Significance was set at Benjamini–Hochberg FDR ≤ 0.10 for metabolites and lipids and FDR ≤ 0.05 for genes. KEGG enrichment of up- and downregulated genes used clusterProfiler^35^ with all tested genes as background (FDR ≤ 0.10, ≥3 genes), and joint pathway analysis of differentially expressed genes and differentially abundant hepatic metabolites used MetaboAnalyst 5.0.^36^

### Paired liver–serum analysis

Of the 188 participants with hepatic and serum polar metabolomics, one principal-component outlier was excluded, leaving 187. Metabolites were matched between compartments by normalized name, retaining annotated features present once on each platform (40 metabolites). Liver–serum agreement was quantified per metabolite by Spearman correlation. Compartment-specific associations with steatosis and lobular inflammation were estimated with limma on the full feature set of each compartment (adjusted for age, sex and BMI), and compartment differences as the effect of histology on the within-participant liver-minus-serum difference after z-scoring each metabolite within compartment. Sensitivity analyses adjusted hepatic models for a triglyceride index (mean of autoscaled triglyceride species) or a sample dilution factor, applied probabilistic quotient normalization, added HbA1c and fasting glucose, or adjusted inflammation for steatosis. Discrimination of steatosis, inflammation, MASH and any fibrosis (stage F1–F4 versus F0) by clinical variables (age, sex, BMI, HbA1c, glucose), serum and hepatic metabolites was estimated by elastic-net logistic regression (glmnet^37^) with 10 × 5-fold stratified cross-validation and compared with DeLong’s test^38^ (pROC^39^). Details are given in the Supplementary methods.

### Partitioned polygenic risk scores

Concordant and discordant pPRS for liver fat^12^ were available for all 211 participants and were standardized within the sample. Their associations with hepatic triglyceride content, log-transformed serum triglycerides, steatosis grade and NAS were estimated by linear regression adjusted for age, sex and BMI. Modification of liver–serum coupling by the discordant pPRS was tested by comparing per-metabolite Spearman correlations above and below its median (2,000 permutations) and by interaction terms. Mediation of pPRS effects on histology through hepatic triglyceride content was estimated with the mediation package^40^ (linear mediator and logistic outcome models adjusted for age, sex and BMI; 2,000 bootstrap resamples).

### External validation of multi-omics signatures in independent cohorts

Transcriptomic signatures were tested in two liver RNA-seq cohorts, GSE135251 and GSE162694,^22,23^ with fibrosis compared as F0 versus F1–F4 and steatosis and inflammation against the NAS; replication required external significance (FDR ≤ 0.05) in the same direction. Hepatic lipid signatures were tested in the ST000915 liver-biopsy lipidomics cohort^10^ (normal versus steatosis). Cohorts, contrasts and analyses are described in the Supplementary methods and **Table S18**.

### Exploratory network and mediation analyses

Weighted co-expression networks^41^ were built for each omics layer, and module–histology and cross-omic module associations were estimated within sex, with formal tests of sex differences and size-matched resampling of women; statistical mediation between transcriptomic and hepatic metabolite modules was tested within sex. These exploratory analyses are described in the Supplementary methods, **Figs. S3–S4 and Tables S13–S16**.

### Statistics and reproducibility

No statistical method was used to predetermine sample size; all participants with available samples were included. Histology was scored blinded to clinical and omics data, and samples were randomized for metabolomics analysis; the analyses were otherwise not blinded. All tests were two-sided, and multiple testing was controlled with the Benjamini–Hochberg method within each analysis. Further information on research design is available in the Nature Portfolio Reporting Summary linked to this article.

## Data Availability

Clinical and matching omics data from individual patients is not available because the inform consent does not cover public deposition of individual patient data. Data can be shared upon request in summary format for all omics features included in the study.
Pooled serum and liver control sample data from the metabolomics study is deposited in MassIVE repository (https://doi.org/doi:10.25345/C50P0X49Q).

## Data availability

Individual-level clinical and omics data cannot be deposited publicly because the informed consent obtained does not cover public release of individual patient data. De-identified summary-level differential-expression, differential-abundance, module-association, pathway-enrichment, mediation, robustness, polygenic risk score and external-validation results are provided in Tables S1–S16 and S19–S20. Analytical conditions and external-dataset descriptions are provided in Tables S17–S18. Additional information is available from the corresponding authors on request. Public datasets analyzed in this study are available from the Gene Expression Omnibus (GSE135251, GSE162694) and the Metabolomics Workbench (ST000915, ST001711). Pooled serum and liver quality-control sample data from the metabolomics study have been deposited at MassIVE and are publicly available as of the date of publication (https://doi.org/10.25345/C50P0X49Q).

## Code availability

The complete R workflow used for data processing, statistical analysis, visualization, and the size-matched sensitivity analysis is publicly available at https://github.com/shashank-KU/masld-sex-stratified-multiomics/tree/main

## Acknowledgements

M.O. and T.H. were supported by the “Investigation of endocrine-disrupting chemicals as contributors to progression of metabolic dysfunction-associated steatotic liver disease” (EDC-MASLD) consortium funded by the Horizon Europe Program of the European Union under Grant Agreement 101136259, and by the Swedish Knowledge Foundation (20220122).

S.R. was supported by the Region Stockholm (ALF project grant, FoUI-1021801), the Swedish Cancerfonden (22 2270 Pj), the Swedish Research Council (Vetenskapsrådet, 2023-02079), the Swedish state under the Agreement between the Swedish government and the county councils (the ALF agreement, ALFGBG-965360), the Swedish Heart Lung Foundation (20220334), the SRP Diabetes Fellowship (2025-00709), the Novo Nordisk Foundation Distinguished Investigator Grant – Endocrinology and Metabolism (NNF23OC0082114), the Novo Nordisk Foundation Project grants in Endocrinology and Metabolism (NNF24OC0091535), a Novo Nordisk donation to the Karolinska Institutet in connection with the professor appointment, and the French government under the France 2030 program (ANR-21-IDES-0006). The European Metropolis of Lille and the University of Lille are thanked for funding and support provided to WILL-CHAIRES-23-001-TREAT.

Views and opinions expressed are those of the authors only and do not necessarily reflect those of the European Union. Neither the European Union nor the granting authority can be held responsible for them. The authors thank Lucas Landmann, Maelle Ullius, and Kattis Kristofferson for assistance in metabolomics analysis.

## Author contributions

Conceptualization and design of study (T.H., S.R., M.O.), clinical study (O.J., R.M.M., U.V.-G., F.T., V.B., D.T., S.R.), metabolomics analysis (D.D., T.H., M.O.), data analysis and interpretation (S.G., with critical input from R.M.M., T.H., S.R., M.O.), manuscript preparation (S.G., T.H., M.O.), critical review and editing (all authors).

