## Supplementary Methods and Figures for "Matched liver–serum multi-omics and genetic risk reveal hepatic compartmentalization of metabolic disease in severe obesity"

#### Sample preparation and mass spectrometry

The samples were randomized and two methods were applied for separate extraction of lipids and polar/semipolar metabolites, and the extracts were then analyzed using an ultra-high-performance liquid chromatography quadrupole time-of-flight mass spectrometry (UHPLC-QTOFMS) as described in Table S17.

Lipidomic analyses were performed on a UHPLC-QTOFMS system (Agilent Technologies, Santa Clara, CA, USA) using a method described previously.[1] An internal standard mixture was used for normalization, and lipid class-specific calibration was used for quantitation. MS data processing was performed using open-source software MZmine 4.3[2] as described below. The identification was done with a custom database, with identification levels 1 and 2 based on the Metabolomics Standards Initiative.[3] However, it should be noted that the method cannot distinguish isomers differing only by their double bond position. Quantification of lipids was performed using a 7-point internal calibration curve (0.1–5 µg/mL). Quality control was performed throughout the dataset by including blanks, pure standard samples, extracted standard samples, and control plasma samples. The RSD of lipids in the pooled plasma samples (n = 9) was on average 16.8%.

The analysis of polar and semipolar metabolites was performed using a method described previously,[4] with an internal standard mixture used for normalization and with calibration curves established for bile acids and amino acids. Identification was based on an in-house spectral library combined with external databases, with identification levels 1 and 2 based on the Metabolomics Standards Initiative. Quality control was performed throughout the dataset by including blanks, pure standard samples, extracted standard samples, and control plasma samples. The RSD of metabolites in the pooled plasma samples (n = 9) was on average 20.5%.

#### Data preprocessing for metabolomics

The raw data were first converted to mzML format using MSConvert in ProteoWizard 3.0.[5] Mass spectrometry data processing was performed using the open-source software package MZmine 4.3. The following steps were applied to the lipidomics data; the polar and semipolar metabolite data were processed with the same steps but with a lower m/z range, an earlier retention-time range and a different noise level: (i) crop filtering with a m/z range of 350–1200 m/z and an RT range of 2.0 to 12 min; (ii) mass detection with a noise level of 750; (iii) chromatogram builder with a minimum time span of 0.08 min, minimum height of 1000 and a m/z tolerance of 0.006 m/z or 10.0 ppm; (iv) chromatogram deconvolution using the local minimum search algorithm with a 70 % chromatographic threshold, 0.05 min minimum RT range, 1000 minimum absolute height, a minimum ratio of peak top/edge of 1.7 and a peak duration range of 0.08–1.0; (v) isotopic peak grouper with a m/z tolerance of 5.0 ppm, RT tolerance of 0.05 min, maximum charge of 2 and with the most intense isotope set as the representative isotope; (vi) join aligner with a m/z tolerance of 0.007 or 7.0 ppm and a weight of 3, a RT tolerance of 0.15 min and a weight of 1 and with no requirement of charge state or ID and no comparison of isotope pattern; (vii) peak list row filter with a minimum of 10 % of the samples; (viii) gap filling using the same RT and m/z range gap filler algorithm with an m/z tolerance of 0.009 m/z or 11.0 ppm; (ix) identification of metabolites was done using a custom database, constructed by analyzing authentic standards with m/z information and retention time information adjusted with internal standards, with an m/z tolerance of 0.009 m/z or 10.0 ppm and a RT tolerance of 0.25 min; and, (x) normalization using the internal standard with closest retention time.

**Network analysis**

Weighted Gene Co-expression Network Analysis (WGCNA)[6] was performed using the blockwiseModules function in the WGCNA R package. Analysis parameters included biweight midcorrelation (corType = 'bicor') to account for outliers, signed correlation networks (TOMtype and networkType = 'signed'), and a sensitive module detection parameter (deepSplit = 4). Module sizes were set to a minimum of 20 features (minModuleSize = 20). Default settings were used for other parameters.

Module eigengenes (MEs) were computed as the first principal component of the expression profiles for each module. The relatedness between modules was examined through eigengene network clustering. MEs were correlated with clinical and molecular variables using Spearman's correlation, and *P* values were adjusted for multiple comparisons using the false discovery rate (FDR).

**Sex differences in module associations**

Module–histology associations (Spearman correlation of each module eigengene with steatosis, lobular inflammation and fibrosis) and cross-omic module–module correlations were estimated separately in men and women. For module–histology associations, *P* values were corrected by the Benjamini–Hochberg method within each omics layer and sex (Table S15a); for cross-omic correlations, between every pair of modules from different omics layers in participants with both layers available, correction was applied across all layer pairs within each sex (Table S15c). Sex differences in each of the 231 module–histology associations were tested by comparing Fisher z-transformed Spearman coefficients between men and women (variance $1.06/[n - 3]$) and, as a sensitivity analysis, by a linear model of the rank-normalized eigengene on the rank-normalized histological feature, sex and their interaction, with Benjamini–Hochberg correction within each omics layer (Table S15b). To separate sex from sample size, women were randomly subsampled 10,000 times to the number of men available in each omics layer or layer pair, the analysis was repeated in each subsample, and the number of significant results (FDR ≤ 0.10) was compared with that in men. Empirical *P* values were computed as $(k + 1)/(B + 1)$ for each direction, and the two-sided *P* value as twice the smaller one-sided value, capped at 1 (Table S15d; Figs. S3c–i and S4).

**Statistical mediation analysis**

Mediation analysis was performed with the mediation R package[7] in participants with all three liver omics layers (n = 186; 50 men, 136 women), stratified by sex. Module eigengenes were scaled to unit variance within each sex, so effects are expressed per standard deviation. To limit the hypothesis space, mediation was tested only for transcriptomic module–mediator module–histology triples in which both modules were themselves associated with that histological feature within sex. Module–trait associations for this screen were computed by Spearman correlation in the complete-case subset on eigengenes scaled to unit variance within sex, with Benjamini–Hochberg correction within each omics layer (transcriptomic and lipid module–trait tests corrected separately). Modules passing FDR ≤ 0.10 were retained, giving 42 candidate triples in men and 235 in women. For each triple, a linear mediator model (mediator ~ transcriptomic module + age + BMI) and a logistic outcome model (dichotomized histological feature ~ transcriptomic module + mediator + age + BMI) were fitted. Models were run in two stages: a quasi-Bayesian approximation (1,000 simulations) across all candidates, with Benjamini–Hochberg correction within sex across the full candidate family; and, for triples at FDR ≤ 0.10, a nonparametric bootstrap (5,000 resamples, bias-corrected and accelerated intervals). Triples with fewer than ten participants in the smaller outcome class were not fitted, and models with fitted probabilities at 0 or 1, or failing to converge, were excluded. Pathways were retained when ACME FDR ≤ 0.10, total-effect FDR ≤ 0.10, and 0 <

proportion mediated ≤ 1; others are reported separately as inconsistent mediation. Sensitivity to sequential ignorability was assessed with medsens on a probit refit of the outcome model, reporting ρ, the mediator–outcome error correlation at which the ACME reaches zero.

**Paired liver–serum analysis**

Participants with both hepatic and serum polar metabolomics were identified after harmonizing sample identifiers. One participant was excluded because of an extreme position in the principal-component space of the metabolomics data (distance from the centroid across PC1–PC5 of 53.9 median absolute deviations, versus 2.1 for the next most extreme participant), leaving 187 participants. Metabolites were matched between compartments by normalized name, retaining only annotated features present exactly once on each platform; ambiguous and unannotated features were excluded, giving 40 matched metabolites. Compartment-specific associations with binarized steatosis and lobular inflammation were estimated with limma[8] on the full feature set of each compartment, adjusted for age, BMI and sex, and then restricted to the matched panel. Because each participant contributed one hepatic and one serum sample, the compartment-by-histology interaction was estimated as the phenotype effect on the within-subject difference (liver minus serum) after z-scoring each metabolite within compartment. This exact paired contrast was the primary analysis. A stacked mixed model using limma's duplicateCorrelation blocked on participant, and an unmoderated ordinary least-squares fit on the same difference, were sensitivity analyses. Agreement between hepatic and serum levels of each metabolite was quantified by Spearman correlation across participants, with Benjamini–Hochberg correction. Directional concordance was assessed by a two-sided sign test on the proportion of metabolites with opposite effect directions, across the full matched panel and within pre-specified bile acid and amino acid classes. FDR ≤ 0.10 was used throughout.

Sensitivity analyses. A hepatic triglyceride (TG) index was calculated for each participant as the mean of the z-scored triglyceride species in the hepatic lipidome. A sample dilution factor was estimated separately for liver and serum on the log2-transformed data before autoscaling: for each feature, the median across the 187 paired participants served as the reference, and the dilution factor of a sample was the median, across features, of its deviation from that reference (the log-scale probabilistic quotient). The base models (age, sex and BMI) were refitted with additional adjustment for the TG index, for the dilution factor of the respective compartment, for HbA1c and fasting glucose, or for the TG index together with HbA1c and glucose; for lobular inflammation, each model was also fitted with steatosis grade as a covariate. In a probabilistic quotient normalization (PQN) model, the hepatic dilution factor was subtracted from the log2 hepatic data before autoscaling. Finally, hepatic polar features correlated with the TG index (|Spearman $\rho$| > 0.5) were removed and the false discovery rate was recalculated (Table S11a). In each model, the paired liver-minus-serum contrast was recomputed with the hepatic data processed as in that model.

Discrimination. Steatosis, lobular inflammation, MASH and any fibrosis (F0 versus F1–F4) were predicted with elastic-net logistic regression (glmnet; $\alpha = 0.5$, $\lambda$ selected as the minimum of the inner five-fold cross-validated deviance). Predictor sets were clinical variables (age, sex, BMI, HbA1c and fasting glucose), serum or hepatic metabolites alone (40 matched metabolites or all features of the compartment), and clinical variables combined with serum or hepatic metabolites; clinical variables were left unpenalized. Missing values were imputed with the training-fold median. Performance was estimated by stratified five-fold cross-validation repeated ten times, and the area under the receiver operating characteristic curve (AUC; pROC) is reported as the mean and range across repeats. Out-of-fold predicted probabilities were averaged across repeats and compared with paired DeLong tests (Table S11e); the halves below and above the median discordant pPRS were compared with unpaired DeLong tests (Table S12g).

**Size-matched sensitivity analysis**

To evaluate whether the larger number of significant module–histology associations observed in women was attributable to unequal sample size and omics availability, we performed 10,000 layer-specific resampling iterations. Women were sampled without replacement to match the number of men with available data separately within serum metabolomics, liver metabolomics, lipidomics, and transcriptomics. The corresponding sample sizes were 57, 52, 57, and 55 participants, respectively. In each iteration, Spearman correlations between module eigengenes and steatosis, inflammation, and fibrosis were recalculated, with false discovery rate adjustment performed separately within each omics layer. Associations with FDR $\leq$ 0.10 were counted and summed across layers. The observed male total was compared with the resulting female resampling distribution using a two-sided empirical probability, $2 \times \min[(k_{\leq} + 1)/(B + 1), (k_{\geq} + 1)/(B + 1)]$, capped at 1. The same procedure was applied to cross-omic module–module correlations, with women matched to the number of men in each layer pair (50–57) and FDR correction across all layer pairs.

### External validation in independent cohorts

Independent validation analyses were performed separately from the discovery analyses, using the external cohorts and datasets summarized in Table S18. For liver transcriptomics, GSE162694 was analyzed using DESeq2 with age and sex as covariates for fibrosis (F0 versus F1–F4), while NAS-based analyses were used to support validation of steatosis and inflammation. GSE135251 was analyzed using the same FDR threshold (0.05), with fibrosis evaluated directly and steatosis and inflammation assessed using NAS. Models for this cohort were fitted without covariate adjustment. A secondary sensitivity analysis of GSE135251 inferred biological sex from expression of sex-specific markers (RPS4Y1, DDX3Y, KDM5D, UTY, and XIST) and incorporated sex into both fibrosis and NAS models. Concordance of log2 fold-change estimates was assessed among genes significant in both the discovery and external datasets using Spearman correlation and directional concordance, with replicated gene- and pathway-level results retained for supplementary reporting. In addition to concordance among genes significant in both datasets, we report the replication rate, defined as the proportion of discovery-significant genes that were also significant and directionally concordant in the external cohort, and genome-wide effect-size concordance.

For liver lipidomics, independent validation was performed using the ST000915 liver biopsy cohort. Steatosis was evaluated using a Normal versus Steatosis comparison, restricted to samples with complete age, BMI, and sex information, with NASH and cirrhosis excluded. The external analysis was adjusted for age, BMI, and sex. Discovery-significant lipid features were assessed for representation in ST000915, directional concordance of effect estimates, and external statistical significance. For EPoS serum metabolomics, the primary validation question was whether discovery-significant features were represented in the external cohort and retained the same direction of association; external statistical significance was considered a complementary measure rather than the sole criterion for validation. The characteristics, references, dataset accessions, and validation contrasts of all external cohorts are summarized in Table S18.

For transcriptomics, validation was assessed using the overlap of genes significant in both the discovery and external datasets, concordance of effect direction, and Spearman correlation of log2 fold-change estimates. For ST000915 liver lipidomics, discovery-significant steatosis-associated lipids were matched to the corresponding external lipid measurements, and agreement was assessed using directional concordance and external statistical significance. For

EPoS serum metabolomics, exact metabolite-name matching was used to determine whether discovery-significant metabolites were represented in the external platform. Directional concordance was evaluated independently of external statistical significance because differences in feature coverage and phenotype definition limited direct replication.

Compact transcriptomic validation results are provided in Table S5, with complete gene-level comparisons in Table S6 and pathway-level validation in Table S7. ST000915 liver-lipidomics results are summarized in Table S8, with complete feature-level comparisons in Table S9. EPoS serum-metabolomics results are summarized in Table S19, with complete feature-level comparisons in Table S20.

**Transcriptomic validation: design and reproducibility**

Discovery DEGs were taken from the MAFALDA DESeq2 analysis of 208 participants, in which genes with ≥10 reads in ≥10 samples were tested for differential expression (21,114 genes; Wald test with independent filtering at $\alpha = 0.05$; Table S1). GSE162694 was analyzed using DESeq2[9] models adjusted for age and sex. Fibrosis was evaluated using a direct F0 versus F1–F4 contrast, whereas steatosis and inflammation were evaluated using NAS as a continuous external phenotype. GSE135251 was used as a second independent liver transcriptomic cohort. Its primary validation analysis used direct fibrosis and NAS models without covariate adjustment. A sensitivity analysis additionally adjusted the GSE135251 fibrosis and NAS models for biological sex. Biological sex was inferred from the expression of Y-chromosome markers, including RPS4Y1, DDX3Y, KDM5D, and UTY, relative to XIST, and was subsequently included as a model covariate. Gene-level validation used FDR ≤ 0.05 for both discovery and external significance filters. Validation was assessed among genes significant in both datasets using the number of shared significant genes, directional concordance, and Spearman correlation of log2 fold-change estimates. Compact numerical summaries are reported in Table S5, and the corresponding gene-level records are provided in Table S6. Among genes significant in both datasets, 46 (GSE162694) and 49 (GSE135251) fibrosis genes were shared with the discovery cohort, with directional concordance of 97.8% and 98.0% and Spearman correlations of log2 fold changes of 0.665 and 0.744. In the NAS-based supporting analyses, 683 and 853 steatosis genes and 761 and 969 inflammation genes were shared, with directional concordance of 98.3–99.4% and Spearman $\rho$ of 0.873–0.894. The only discordant fibrosis gene in either cohort was KRT10. Expressed as replication rates (significant and directionally concordant externally), these corresponded to 48–52% of fibrosis, 44–55% of steatosis and 41–52% of inflammation discovery genes, and genome-wide concordance was lower (Spearman ρ 0.34–0.57). After adjustment for inferred sex in GSE135251, 50, 868 and 981 shared genes were identified for fibrosis, steatosis and inflammation, with directional concordance of 98.0–98.5% and Spearman ρ of 0.768–0.892, indicating that external replication was not attributable to sex imbalance in that cohort.

**Pathway-level transcriptomic validation**

To extend validation beyond individual genes, upregulated and downregulated gene sets were analyzed separately using KEGG pathway enrichment. Pathways were retained at Benjamini–Hochberg FDR ≤ 0.10 with at least three genes, and a pathway was considered replicated only when it met these criteria in both the discovery and the external dataset. The discovery enrichment used all tested genes as background. Of seven pathways enriched among discovery fibrosis DEGs, six were enriched in at least one external cohort and one (bladder cancer) in both. The complete pathway-level results, including pathway identifiers, descriptions, enrichment statistics, direction of association, cohort, phenotype, and contributing gene identifiers, are provided in Table S7. The corresponding gene-level validation records are provided in Table S6, allowing pathway-level agreement to be traced to the contributing genes. Pathway-level replication can be influenced by differences in cohort composition, library preparation, gene annotation, and the phenotype definitions used in the external datasets. Therefore, pathway agreement was interpreted as supportive biological replication rather than as an exact reproduction of the discovery enrichment landscape.

**ST000915 liver lipidomics: design, feature matching, and interpretation**

Discovery-stage steatosis-associated lipid signatures were evaluated in the independent ST000915 liver-biopsy lipidomics cohort. Histologically normal samples were compared with samples with steatosis; samples classified as NASH or cirrhosis were excluded, and the analysis was restricted to samples with complete data on age, BMI and sex (29 normal, 17 steatosis). External associations were adjusted for age, BMI and sex. Of the 854 steatosis-associated lipid features in the discovery cohort (FDR ≤ 0.10), 187 were annotated, and 68 measurements corresponding to 54 unique species were present in ST000915. Of these 68, 51 (75.0%) showed the same direction of association in both cohorts, and 49 were significant externally (FDR ≤ 0.10). Of the 49, 47 were triglycerides concordant with the discovery direction. The two significant discordant lipids were TG(56:6) and CE(18:1), both decreased with steatosis in the discovery cohort and increased in ST000915. Concordance was strongly class-dependent: 48 of 50 triglyceride measurements were concordant, compared with 3 of 18 membrane and neutral-ester lipids, and 13 of 14 phosphatidylcholine measurements showed the opposite direction. After de-duplication of repeated identifiers, concordance was 77.8% (42 of 54 unique species). External support is therefore limited to the triglyceride component of the steatosis signature, and it is further constrained by the limited overlap in lipid coverage between platforms. Summary and feature-level results are given in Tables S8 and S9.

**EPoS serum metabolomics: design, feature matching, and interpretation**

Because only three of the 16 discovery serum associations were represented in EPoS, this comparison is reported in the Supplementary Information only. The EPoS polar-metabolomics dataset[1] contained 36 named metabolites measured across 627 participants. The downloaded metabolomics data were already log2-transformed and scaled to zero mean and unit variance. Therefore, no additional normalization or scaling was applied during external validation. The downloaded external metadata did not provide a consistently usable grade-0 category. Phenotype-specific validation comparisons were therefore used rather than attempting to reconstruct the discovery contrasts exactly. The external comparisons were grade 1 versus grades 2–3 for steatosis, grade 1 versus grades 2–3 for inflammation, and stage 1 versus stages 2–4 for fibrosis. At the discovery level, 12 steatosis-associated metabolites and four inflammation-associated metabolites met FDR $\leq$ 0.10. No fibrosis-associated serum metabolites met the discovery significance threshold. Exact-name matching to the EPoS metabolomics panel identified two of the 12 steatosis-associated metabolites and one of the four inflammation-associated metabolites. Both steatosis-associated metabolites represented in EPoS, glutamic acid and leucine/isoleucine (matched to EPoS isoleucine), showed the same direction of association as in the discovery serum cohort, although neither reached external significance at FDR $\leq$ 0.10. For inflammation, leucine/isoleucine was the only discovery-significant metabolite represented in EPoS. The external inflammation association was in the opposite direction and was not statistically significant. Because no discovery-significant serum metabolites were identified for fibrosis, feature-level fibrosis validation was not possible.

The phenotype-level validation results are summarized in Table S19, and the complete matched and unmatched feature-level records are provided in Table S20. These findings illustrate why external FDR significance alone was not used as the primary validation criterion. For steatosis, neither represented metabolite reached external FDR $\leq$ 0.10, but both retained the same direction of association as in the discovery cohort. Conversely, six EPoS metabolites were associated with inflammation at FDR ≤ 0.10, but none was among the discovery-significant features; the only discovery-significant inflammation metabolite represented in EPoS (leucine/isoleucine, matched to isoleucine) was not significant externally (FDR = 0.97) and showed the opposite direction. External-significant feature counts and directional concordance therefore provide complementary information and should not be interpreted interchangeably. Overall, the EPoS serum-metabolomics analysis provided limited directional support for selected annotated discovery metabolites. Incomplete feature coverage, differences in phenotype definition, and the absence of a consistently usable grade-0 category limited direct replication.

**Fig. S1.** Principal component analysis (PCA) of liver samples. PCA plot of variance-stabilized transformed (VST) counts from liver samples, colored by sex (women and men). The PCA was performed using DESeq2. The plot shows the first two principal components (PC1 and PC2), which explain 21% and 8% of the variance, respectively. Each point represents a sample, with ellipses indicating the 95% data ellipses for each sex group. Filled points are color-coded by sex. The analysis reveals distinct clustering of samples based on sex, suggesting sex-specific differences in liver gene expression profiles.

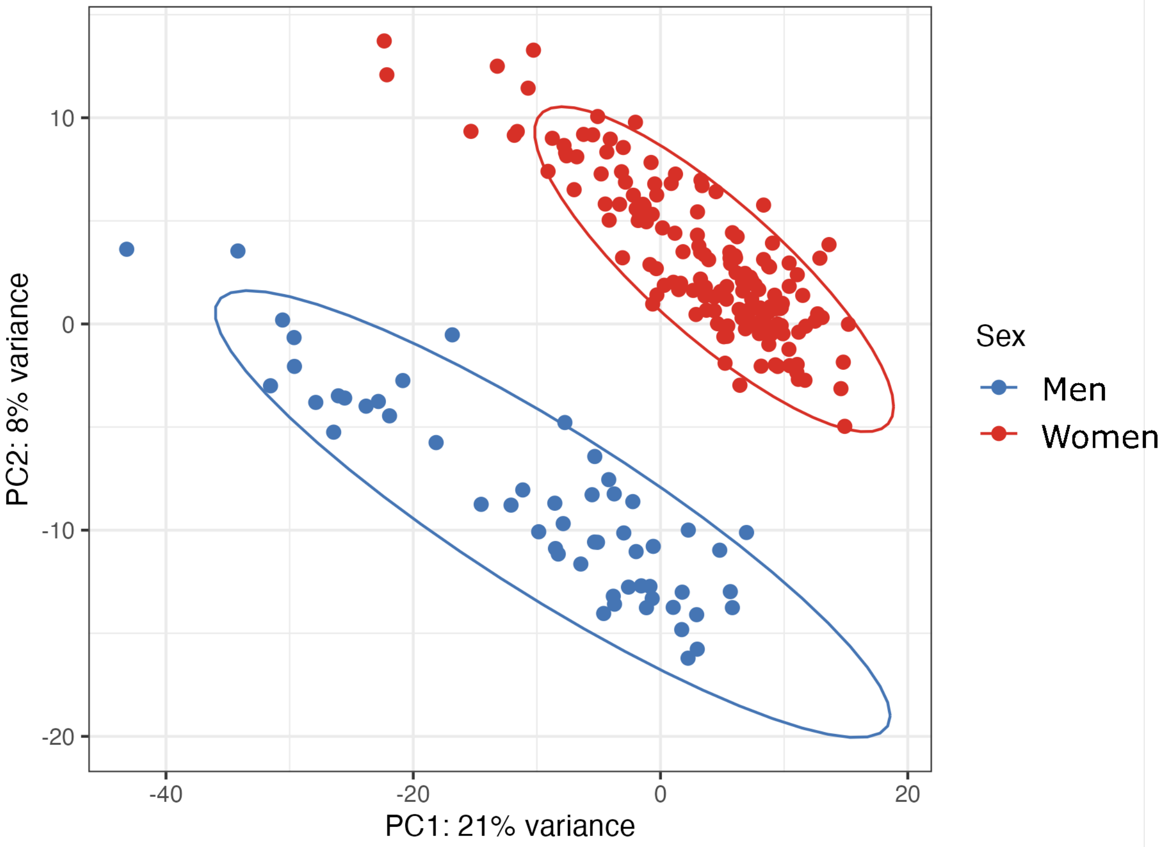

**Fig. S2. Joint pathway analysis of histology-associated hepatic genes and metabolites.** MetaboAnalyst 5.0 joint pathway analysis of differentially expressed genes (FDR ≤ 0.05; Table S1) and differentially abundant hepatic metabolites (FDR ≤ 0.10; Table S4) associated with steatosis (blue) and lobular inflammation (red). Each dot is a metabolic pathway; x axis, number of hits (matched genes and metabolites) in the pathway; dot size, pathway impact (topology-based importance of the matched nodes within the pathway).

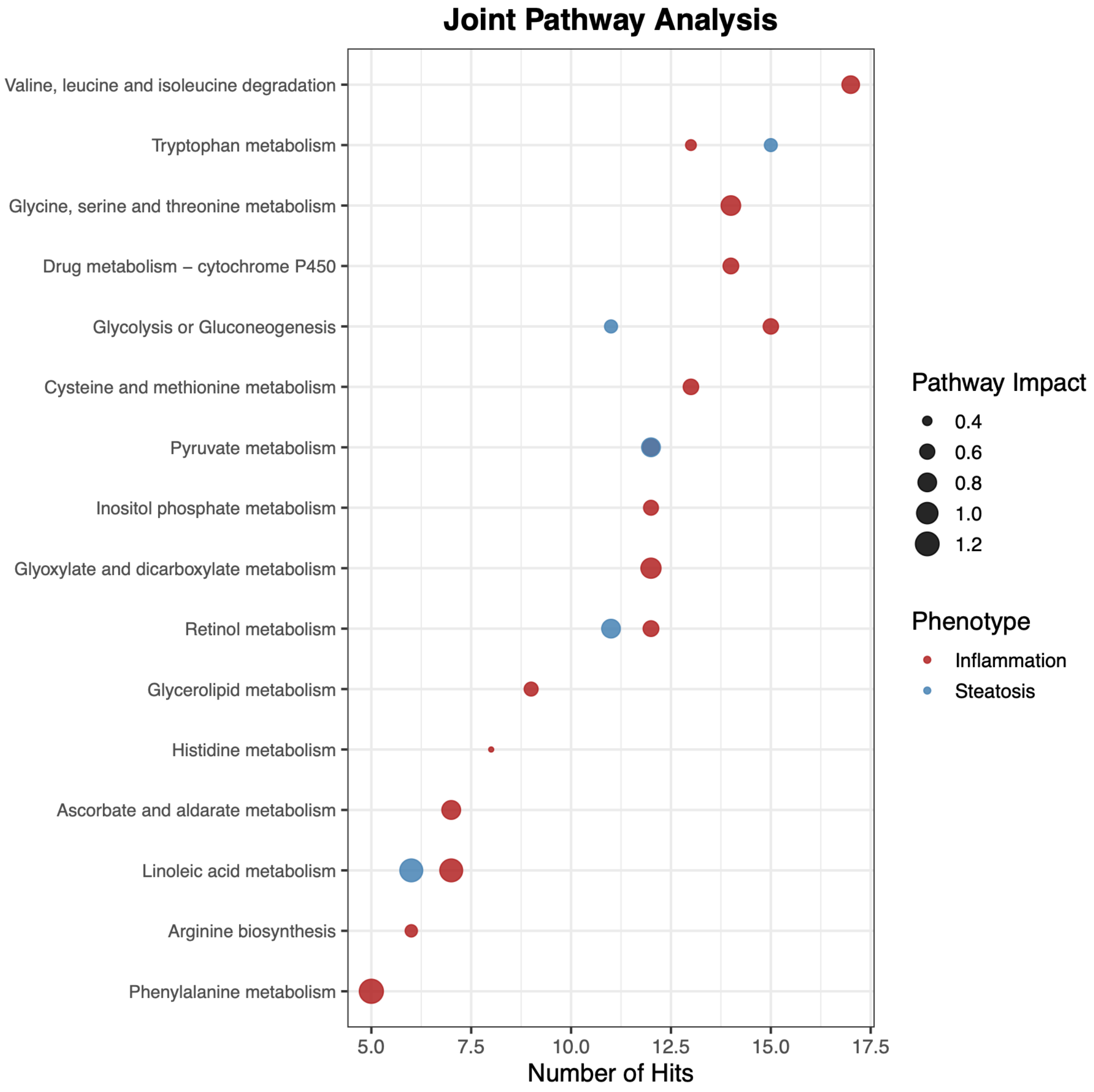

**Fig. S3. Cross-omic module–module correlations in men and women.** (a, b) Correlations between module eigengenes of different omics layers at FDR ≤ 0.10 (Spearman; Benjamini–Hochberg across all layer pairs) in men (a; 59 correlations, n = 50–57) and women (b; 311 correlations, n = 135–153). Sectors are modules, colored by layer; orange, positive; blue, negative; opacity is proportional to |ρ|. (c–i) Size-matched subsampling of women: 10,000 subsamples matched to the number of men in each layer pair. Histograms show the number of significant cross-omic correlations for all layer pairs combined (c) and for each pair (d–i); dashed line, count in men; two-sided empirical *P* values (Table S15d).

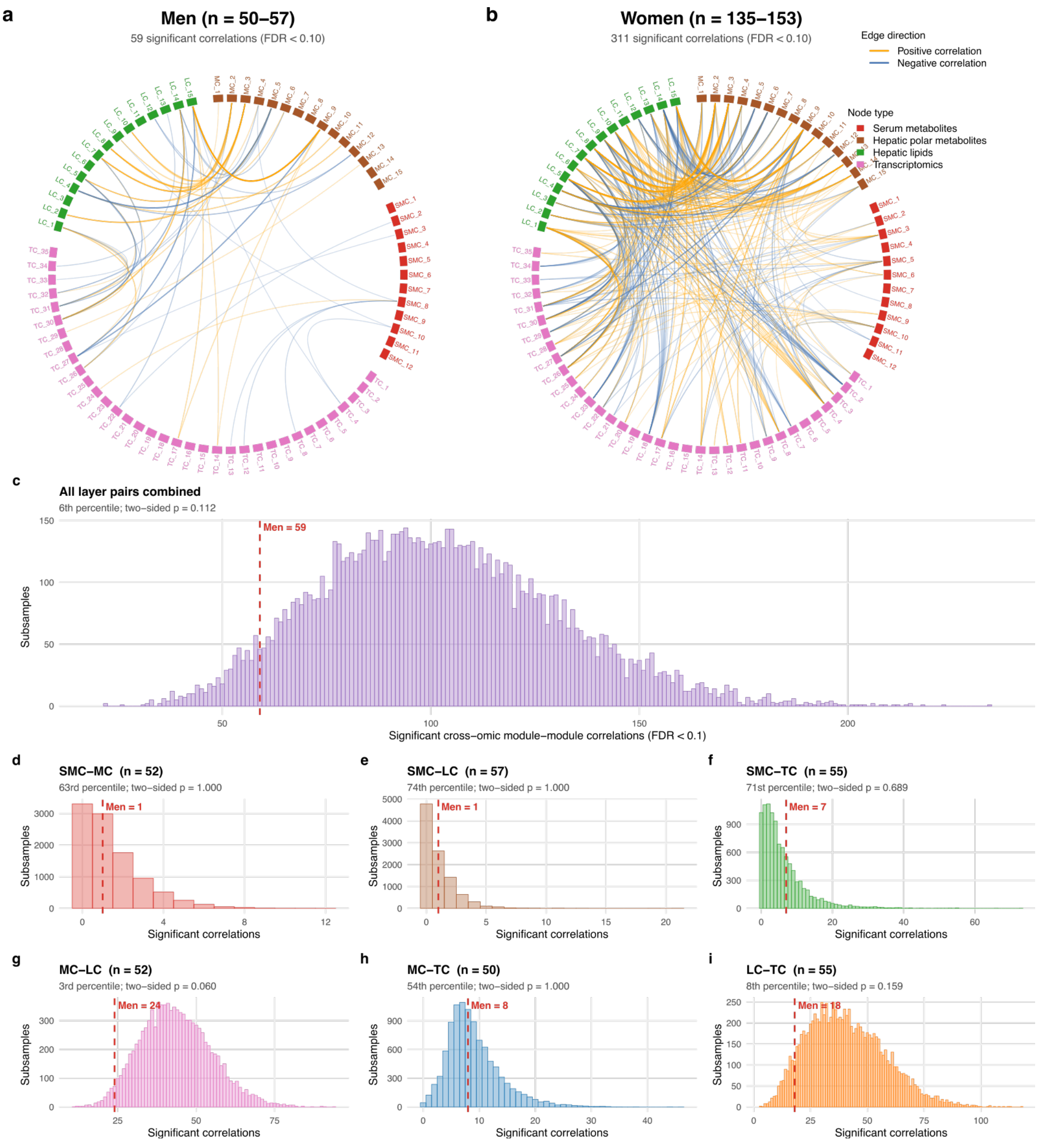

**Fig. S4. Size-matched subsampling of women for module–trait associations.** (a) Distribution of the total number of significant module–trait associations (FDR ≤ 0.10), summed across the four omics layers, in 10,000 random subsamples of women, each matched within each layer to the number of men with data in that layer (serum metabolomics 57, hepatic polar metabolomics 52, hepatic lipidomics 57, transcriptomics 55). The dashed red line marks the count observed in men (21), which lay at the 41st percentile of the size-matched distribution (two-sided *P* = 0.82); the larger number of associations in all women (51; n = 152–154) is therefore consistent with their larger sample size. (b–e) The same subsamples by omics layer: serum metabolomics (b; men 0; *P* = 1.00), hepatic polar metabolomics (c; men 0; *P* = 1.00), hepatic lipidomics (d; men 16; *P* = 0.65) and transcriptomics (e; men 5; *P* = 0.60). Empirical *P* values are two-sided, computed as twice the smaller of the one-sided values (k + 1)/(B + 1), capped at 1 (Supplementary methods; Table S15d).

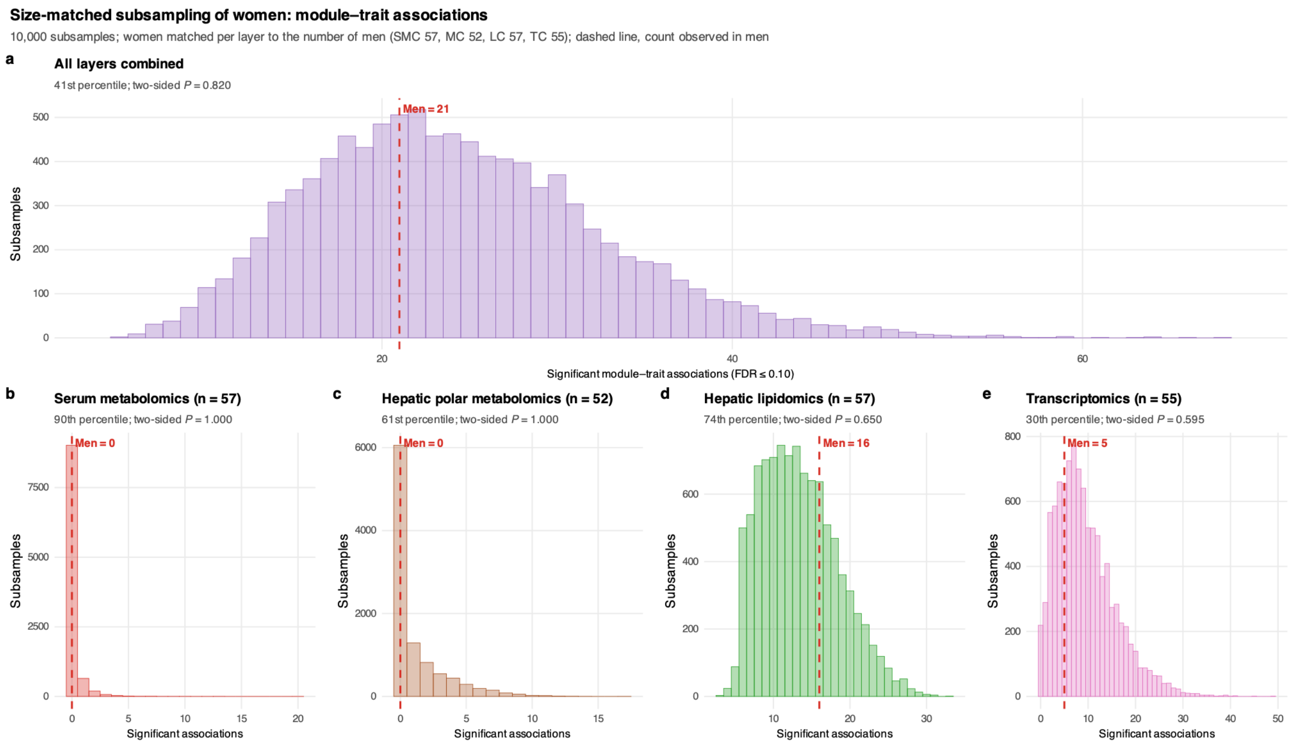

**Fig. S5. Study design and summary of findings.** (a) Adults with severe obesity (n = 211; 154 women, 57 men) underwent bariatric surgery with protocol liver biopsy; liver tissue and serum were obtained in the same operative session, so liver–serum differences are not attributable to differences in fasting state, time of day or sampling interval. Steatosis, lobular inflammation and fibrosis (F0 versus F1–F4) were graded with the Kleiner system. (b) Molecular layers, number of participants and number of features; the matched panel comprised 187 participants and 40 metabolites measured once on both platforms (12 amino acids, 10 bile acids and 18 other metabolites). (c) Liver–serum Spearman correlation (across participants) of the 40 matched metabolites (box, median and interquartile range; median ρ 0.02, interquartile range −0.06 to 0.14; filled points, FDR ≤ 0.10, 9 of 40; Table S10b). (d) Polar features associated with steatosis and lobular inflammation (limma adjusted for age, sex and BMI; FDR ≤ 0.10) in liver and serum of the matched subset (n = 187; Table S11a) and in serum of all participants with serum data (n = 209; Table S10a). (e) Effects of lobular inflammation on the 12 matched amino acids and 10 bile acids in liver and serum (standardized effect, SD units). Dark purple, the six amino acids with a significant liver–serum difference (FDR ≤ 0.10); 9 of 10 bile acids had opposite liver–serum directions (two-sided sign test $P$ = 0.021; Table S10b). The amino acid discordance persisted after adjustment for a hepatic triglyceride index, sample dilution, probabilistic quotient normalization, or HbA1c and glucose (hepatic associations 75–291 across phenotypes and models; all six amino acids retained for inflammation), but only lysine and valine remained after adjustment for steatosis (Table S11a). (f) Cross-validated AUC (elastic net, 10 × 5-fold) of clinical variables (age, sex, BMI, HbA1c and glucose) alone or with serum or hepatic metabolites (Table S11d); adding serum improved discrimination of steatosis by 0.02 (DeLong $P$ = 0.03; Table S11e). (g) Associations of the discordant and concordant partitioned polygenic risk scores (pPRS) with hepatic and serum triglycerides (TG), steatosis grade and NAS (β per SD with 95% CI; linear regression adjusted for age, sex and BMI; n = 211; Table S12a); the same discordant score was associated with higher hepatic and lower serum TG. (h) Proportion of the discordant pPRS effect on lobular inflammation and MASH mediated by hepatic TG content (2,000 bootstrap resamples; Table S12b); mediation of steatosis was effectively complete (102%) and is not shown. (i) External validation of differentially expressed genes in GSE162694 and GSE135251 (Table S5) and of steatosis-associated triglyceride and phosphatidylcholine species in ST000915 (29 normal versus 17 steatotic biopsies; Tables S8–S9). Filled points, same direction; open points, replicated (significant in both cohorts in the same direction); labels, number in the same direction /

number replicated. Significance thresholds were FDR ≤ 0.05 for genes and FDR ≤ 0.10 for metabolites and lipids. AUC, area under the receiver operating characteristic curve; FDR, false discovery rate; MASH, metabolic dysfunction-associated steatohepatitis; NAS, NAFLD activity score; TG, triglycerides.

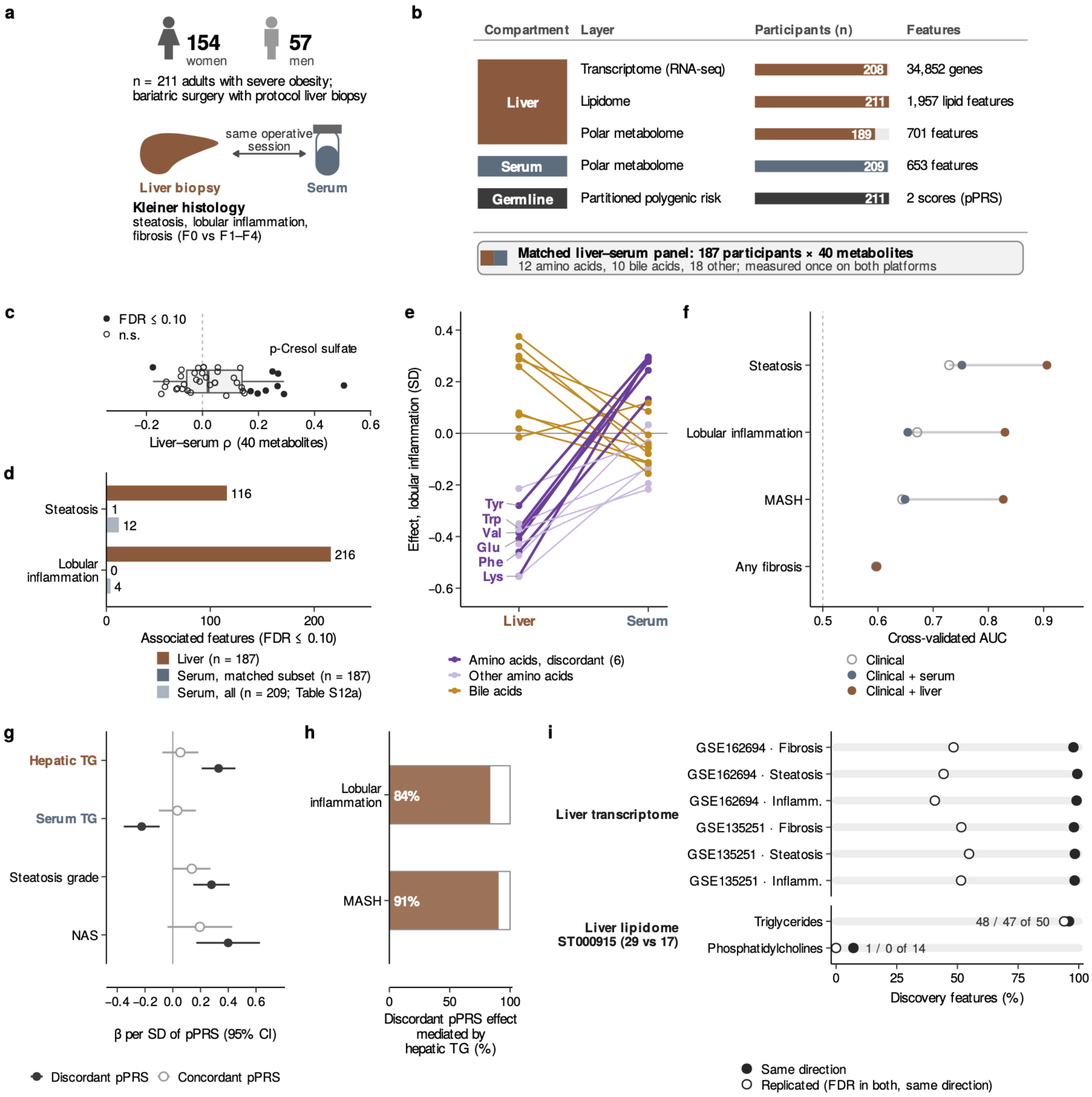
